# Language models reflect clinical evidence but fail to adapt it to patients

**DOI:** 10.64898/2026.09.14.26363047

**Authors:** Shuhan He, Joshua W. Joseph, Pedram Safari, Allison Goff, Pawel Jan Slusarz, Amal Mohamed, Spencer Lord, Joshua N. Goldstein, Ali S. Raja

## Abstract

**Background:** Clinical language models must use evidence to produce the number and action required by a particular case. Whether numerical knowledge reliably becomes a correct clinical response is unclear.

**Methods:** NUMBERS evaluated 16 model configurations on 1,300 questions linked to public clinical evidence. Linked experiments tested prevalence updating, patient-specific estimates and clinical actions. The direct-action experiment compared cutoff recall plus action selection with a supplied complete rule across 50 rules, five models and 7,500 calls.

**Results:** Among diagnostic estimates outside the source-result tolerance, 77.1% remained within the evidence’s central 80% predictive range. Models updated prevalence-dependent quantities correctly in 78.5% of comparisons with inputs and a calculation request, versus 33.7% with clinical wording. Supplying inputs and requesting calculation raised patient-level near-target answers from 29.7% to 78.8% across 2,340 pairs. Models selected an incorrect action despite stating a cutoff that implied the correct action in 413 of 3,750 recall-arm calls (11.01%; 95% confidence interval, 8.93 to 13.17). Supplying the complete rule raised action accuracy from 85.63% to 99.41%, an improvement of 13.79 percentage points (95% confidence interval, 11.65 to 15.95).

**Conclusions:** Models often produced evidence-consistent numbers but failed to adapt them to a case or act consistently with their own stated cutoff. Explicit inputs, calculations and complete rules substantially improved performance in controlled prompts.

**Funding:** National Academy of Medicine, Agreement No. 2026A008797

## Introduction

Clinical numbers only make sense in context. The meaning of a positive test depends on disease prevalence; treatment benefit depends on baseline risk; prognosis depends on who was studied and for how long. As language models are proposed for increasingly autonomous clinical work,[1] they must do more than recall a plausible number. They must use the evidence for the population or patient described.[2,3,4]

Evaluations test medical knowledge,[5,6] simulated patient interaction,[7] broader clinical tasks[8,9] and numerical answers or calculations.[10,11] Calls to move beyond medical examinations[12] and evaluate models across tasks[13] raise a specific question: can a model use what it knows when the case changes? Numerical relationships make that question testable. Changing prevalence must change a predictive value, whereas changing a patient’s measurement must leave an authoritative cutoff fixed. We report these tests using TRIPOD-LLM.[14] Models can reason through calculations or use external tools,[15,16,17,18] but those capabilities help only when the system recognizes what the clinical question requires.

NUMBERS (Numerical Understanding in Medicine: Benefits, Effects, Risks, and Statistics) follows evidence through population, patient and action. We tested estimates against published results and the surrounding evidence, then asked whether models updated quantities for a changed population or patient and held clinical rules fixed as case values changed. The direct-action experiment tested the final link: whether a model chose the action implied by its own stated cutoff, and whether supplying the complete rule improved the choice.

## Methods

### Design and clinical evidence

The core benchmark comprised 1,300 questions from 847 source studies or database records, with 5,558 reference values covering 26 measures in five estimation families. Sources included diagnostic meta-analysis datasets, national surveys and CDC public data, and structured ClinicalTrials.gov results.[19,20,21,22,23,24,25,26] Questions retained clinical context while withholding the numerical result and calculation inputs. Sixteen configurations answered under two response instructions, three times each: 124,800 calls and 533,568 quantity requests. No model was trained or tuned; calls used separate conversations without retrieval, browsing or calculation tools. Model identifiers and settings are reported in Supplementary Tables S4 and S42.

Each experiment was frozen before its own calls; later experiments were designed after earlier results were known. The prevalence-update extension was non-confirmatory. Patient-level and original action-level primary outcomes were prespecified. Supplementary Table S42 records their designs and analysis status.

### Numerical and patient-level outcomes

Source-result accuracy meant falling within the prespecified error allowance: 0.05 for probabilities, log(1.5) for positive ratios, and measure-specific allowances for other quantities. A very-large error exceeded twice the allowance. Refusals, invalid responses and missing requests remained in all-request denominators. Secondary diagnostic analyses compared valid answers with the predictive distribution obtained by refitting the other studies in each meta-analysis.[27,28,29]

The prevalence-update experiment used 100 diagnostic items and 14 configurations. Its 2,300 eligible model-item-quantity comparisons used each model’s median baseline estimate to define the required update. Matched prompts described the prevalence change clinically, supplied inputs and requested the calculation, or restated unchanged prevalence with an irrelevant detail. Correct updates were within 0.05 of the required value; clinical updates also had to move in the required direction.

The patient-level experiment used 200 diagnostic or survey-derived items,[22,30] 13 configurations and 64,740 scheduled calls across its modules. Matched group and patient prompts described old and changed values. Near-target answers were within 0.05 of the changed value and closer to it than to the old value. Three primary outcomes measured movement toward that target, patient versus group wording, and the share of the required change carried. A prespecified secondary experiment supplied inputs and requested calculation. Empirical targets were prevalences of the described subgroup, not measurements of an individual.

### Clinical-action experiments

The original experiment evaluated 50 official NICE, USPSTF and CDC recommendations with ten model versions, five values around each cutoff, three prompt formats, two instructions and three repetitions: 45,000 responses. Actions were inferred from the reported cutoff and source comparator. The prespecified primary contrast compared critical action errors near or at the cutoff with those farther away; sequence analyses measured cutoff variation and wrong-direction reversals.

The direct-action experiment explicitly requested an action. Five models answered five case values for each of 50 rules, in three repetitions, under two matched conditions: recall the cutoff and select an action, or select an action with the complete cutoff, comparator and action map supplied. This produced 7,500 calls, 3,750 per condition. Strict discordance required a valid stated cutoff whose implied action was correct under the source comparator, alongside an explicitly selected incorrect action; its denominator was all scheduled recall-arm calls. Accuracy counted all scheduled calls in each condition. Paired analyses distinguished valid wrong-to-right changes from recovery of invalid responses.

Collection proceeded through 12, 24 and 14-rule groups and finished on August 30, 2026; the prespecified 24-rule confirmation retained its primary analysis status, while the overall estimate described performance across all 50 rules. Supplementary Methods section 6 and Table S67 report that analysis, the progression rules and a sensitivity analysis including eight earlier development rules excluded from the main estimate.

### Statistical analysis and oversight

Intervals generally used 10,000 bootstrap resamples with observations from each source kept together.[31] Patient-level estimates summarized equally weighted configuration-by-module cells; Holm correction covered the three primary tests.[32] The original action experiment used recommendation-level resampling and sign-flip permutation tests. Direct-action estimates gave each rule equal weight and used 100,000 rule-level bootstrap resamples; intervals describe rule sampling for the fixed model panel. Exploratory tests and the development-rule sensitivity are identified separately. Full scoring, multiplicity rules[33] and response accounting appear in the Supplementary Appendix.

Mass General Brigham determined that the public-data study was not human-subjects research (REDCap 4737; August 14, 2026). No patients were enrolled and no care was delivered.

## Results

### Models reflected the surrounding clinical evidence

Under the best-estimate instruction, source-result accuracy ranged from 28.9% for diagnostic performance to 77.8% for time-to-event quantities. No configuration led every estimation family. Positive predictive value was especially difficult: 18.5% of requests were accurate and 54.5% had very-large errors (Supplementary Figure 24).

Many source-result misses were nevertheless compatible with the evidence. In one dementia-screening question, the withheld study reported sensitivity of 0.65 and a model answered 0.85. The other studies supported a predictive range of 0.57 to 0.92. Across 109,897 valid diagnostic answers, 74,349 missed the source-result tolerance; 77.1% of those misses (95% confidence interval [CI], 73.0 to 80.2) lay inside the central 80% predictive range. Accuracy tracked what the surrounding studies could predict across 24 clinical-area-by-measure cells (Spearman rho, 0.77; exploratory P=0.000011; Figure 1).

**Figure 1.**
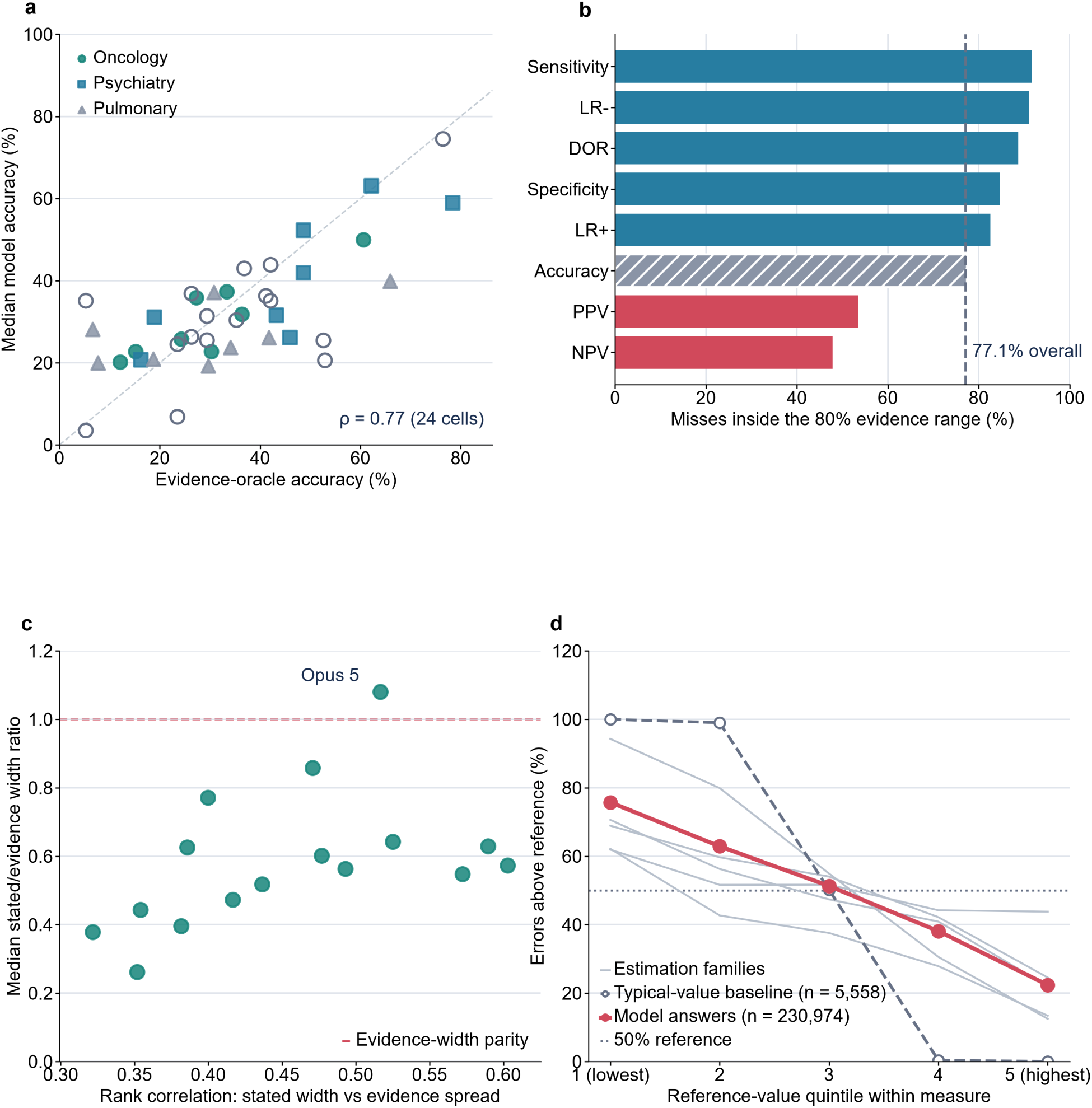
| Evidence-consistent estimates can still miss the population-specific quantity. **a**, Median accuracy across 16 configurations versus accuracy of a model-independent evidence estimate, across 24 clinical-area-by-measure cells. Circles, squares and triangles identify oncology, psychiatry and pulmonary medicine. Hollow symbols show neurology and cardiology below the support threshold and excluded from the correlation (Spearman rho, 0.77; exploratory P=0.000011). **b,** Share of 74,349 valid diagnostic source-result misses inside the central 80% predictive range; the dashed line marks 77.1% overall. **c,** Each configuration’s stated-range correlation with evidence spread and median stated-to-evidence width ratio; parity is marked. **d,** Errors above the reference by within-measure reference quintile: 230,974 model answers and a source-held-out typical-value baseline from 5,558 references. Thin lines show estimation families. Panels b and d are post hoc. LR, likelihood ratio; DOR, diagnostic odds ratio; PPV and NPV, positive and negative predictive values.

The pattern was consistent with learned numerical regularities. A typical-value baseline was accurate for 81.0% of hazard ratios, versus 77.8% for the models. Atypical values were pulled toward typical values in 23 of 26 measures. Evidence consistency was weaker for predictive values, which depend on prevalence: only 53.4% of positive-predictive-value misses and 47.9% of negative-predictive-value misses fell inside the evidence range. A plausible literature value could therefore miss the population-specific quantity.

### Explicit calculation repaired population updating

For AUDIT-C, an alcohol-screening questionnaire, the model’s baseline positive predictive value was 0.50 at a prevalence of 36.6%. Updating from that starting estimate when prevalence halved to 18.3% gave a target of 0.28. With clinical wording the model answered 0.502; with inputs and a calculation request it answered 0.280 (Figure 2).

**Figure 2.**
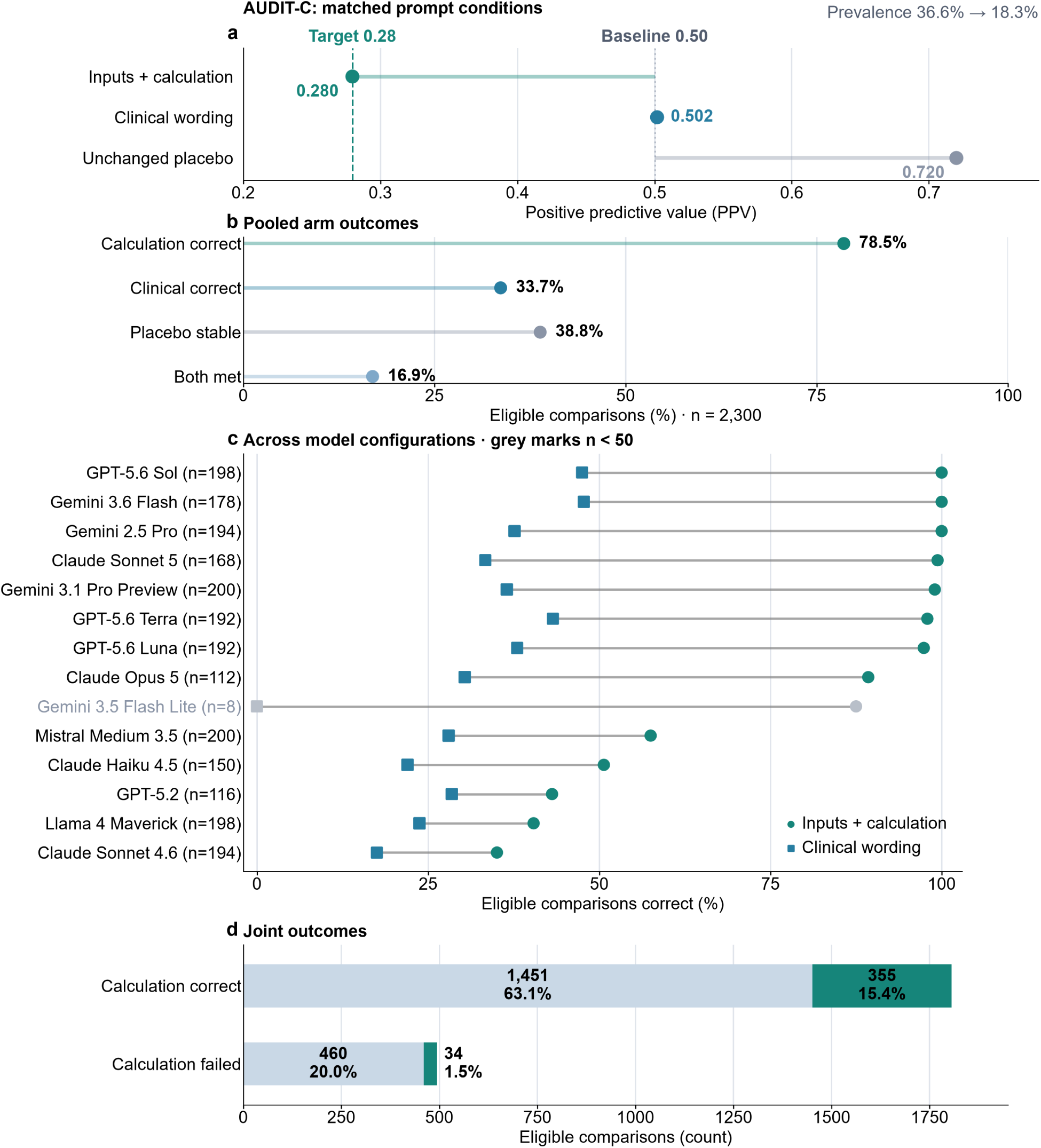
| Explicit calculation improves population updating. **a**, GPT-5.6 Sol: a baseline estimate of 0.50 at 36.6% prevalence implies 0.28 at 18.3%. This model-anchored target differs from Figure 3’s source-based target. **b,** Outcomes among 2,300 eligible comparisons; both conditions means a correct clinical update and unchanged placebo. **c,** Accuracy by configuration: circles, calculation; squares, clinical wording; gray, fewer than 50 comparisons. **d,** Joint outcomes: green denotes both conditions, pale blue their absence. Percentages use all 2,300 comparisons. The calculation advantage was 44.9 percentage points (95% source-cluster bootstrap interval, 39.0–50.9; exploratory P<0.001).

Across 2,300 eligible comparisons, updates were correct in 78.5% with inputs and a calculation request, versus 33.7% with clinical wording (difference, 44.9 percentage points; 95% CI, 39.0 to 50.9; exploratory P<0.001). The same arithmetic framed as testing manufactured parts reached the target in 97.8% of answers. Calculation capability did not reliably become a correct clinically framed update.

A correct clinical update accompanied by a stable answer to redundant context occurred in 16.9% of comparisons. Of 1,806 comparisons answered correctly when calculation was requested, 1,451 (80.3%) lacked that combination. The redundant-context answer moved by more than 0.05 or became unusable in 61.2% (95% CI, 55.7 to 66.9). Further analyses implicated the restated prevalence rather than the irrelevant scheduling detail (Supplementary Figure 11).

### Patient information moved answers without reliably reaching the target

Patient-level answers were 31.0 percentage points more likely to be near the changed value than near the old value (95% CI, 20.6 to 39.2; Holm-adjusted P=0.0006). Patient versus matched group wording showed a difference of 0.0 points (95% CI, −2.0 to 1.6; adjusted P=0.98). Models carried a median 81.4% of the required change (95% CI, 75.5 to 86.8; adjusted P=0.0006 against the 50% reference). These were the three prespecified primary outcomes (Table 1).

**Table 1.** | Principal findings across evidence, population, patient and action. Intervals use source or rule resampling for fixed model panels. Patient primary estimates summarize configuration-by-module cells; the repair median and pooled percentages are different summaries. Direct discordance is an incorrect selected action despite a stated cutoff implying the correct action under the source comparator. The overall 50-rule estimate excludes eight development rules; the prespecified 24-rule confirmation and collection groups are reported in Table S67. Scheduled denominators include invalid responses. Near-target means within 0.05 of the changed value and closer to it than to the old value.

| Assessment and analysis status | Analysis set | Result (95% confidence interval) |
| --- | --- | --- |
| Evidence consistency (secondary) | 74,349 diagnostic source-result misses | 77.1% inside the central 80% evidence range (73.0–80.2) |
| Population update (non-confirmatory) | 2,300 eligible comparisons; 14 configurations | 78.5% inputs + calculation; 33.7% clinical wording; difference 44.9 points (39.0–50.9), exploratory |
| Patient target versus old value (primary) | 26 configuration-by-module cells | Difference 31.0 points (20.6–39.2); Holm-adjusted P=0.0006 |
| Patient versus group wording (primary) | 25 estimable paired cells | Difference 0.0 points (–2.0 to 1.6); Holm-adjusted P=0.98 |
| Required patient-level change carried (primary) | 25 estimable paired cells | Median 81.4% (75.5–86.8); Holm-adjusted P=0.0006 against 50% |
| Patient calculation repair (secondary) | 2,340 scheduled pairs; 13 configurations | Near-target: 29.7% → 78.8% pooled; median configuration gain 58.9 points (45.3–69.6) |
| Inferred-action errors near versus far (primary) | 45,000 responses; 50 rules; ten models | 8.47% versus 6.56%; difference 1.91 points (0.40–3.55); P=0.011 |
| Direct action: overall 50-rule estimate | 50 rules; five models; 3,750 calls per arm | Discordance 11.01% (8.93–13.17); accuracy 85.63% → 99.41%; gain 13.79 points (11.65–15.95) |

The AUDIT-C example connects movement with the remaining error. The population task tested updating from the model’s own starting estimate; the patient task tested the value calculated from source inputs. Sensitivity and specificity of 0.75 imply a positive predictive value of 0.633 at a pretest probability of 36.6%, and 0.401 at 18.3%. The model moved from approximately 0.69 to 0.47 when given the changed patient information; with the inputs and a calculation request it returned 0.401 (Figure 3).

**Figure 3.**
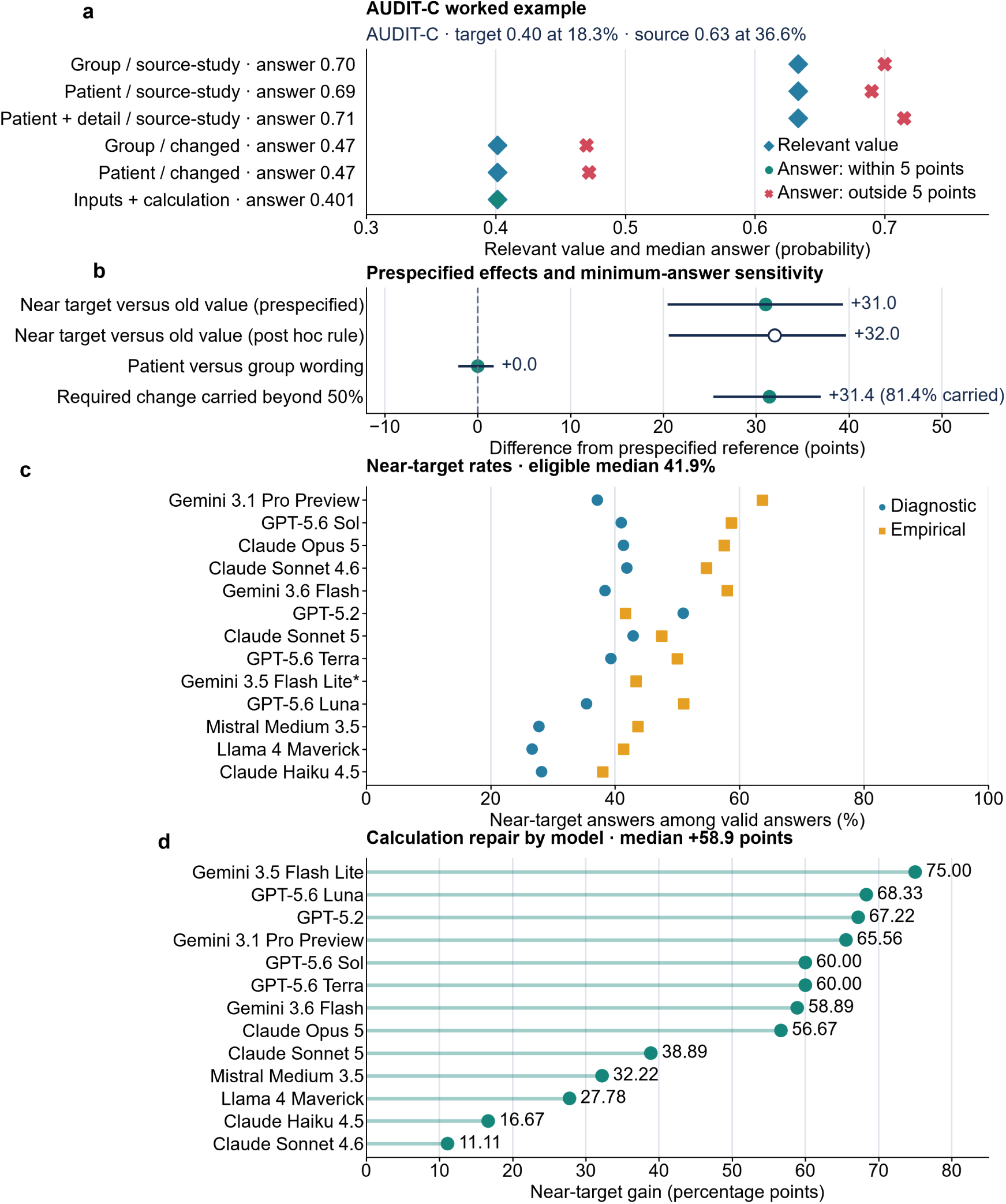
| Patient information moves estimates; calculation improves accuracy. **a**, Six GPT-5.6 Sol AUDIT-C conversations; sensitivity and specificity of 0.75 imply targets of 0.633 and 0.401. **b,** Prespecified effects with 95% source-cluster bootstrap intervals; references are zero for the first two outcomes and 50% for change carried. The hollow point applies the post hoc minimum-answer rule. **c,** Near-target rates among valid answers. The asterisk marks the single-answer diagnostic cell excluded from the 41.9% eligible-cell median (counts: Table S37). **d,** All 2,340 repair pairs, 180 per configuration; invalid answers count as not near-target. Median improvement: 58.9 percentage points (95% interval, 45.3–69.6; unadjusted secondary analysis).

Across 2,340 scheduled repair pairs, near-target answers increased from 29.7% to 78.8%. The median improvement across 13 configurations was 58.9 percentage points (95% CI, 45.3 to 69.6; unadjusted secondary interval). Pooled percentages and the median configuration improvement summarize different quantities. Every configuration improved.

### Models selected actions inconsistent with what their cutoffs implied

NICE recommends platelet transfusion in active upper gastrointestinal bleeding when the count is below 50 × 10⁹/L.[34] At a count of 60 × 10⁹/L, one model stated the cutoff of 50 but selected platelet transfusion. Its own stated cutoff implied the correct alternative under the published comparator (Figure 4).

**Figure 4.**
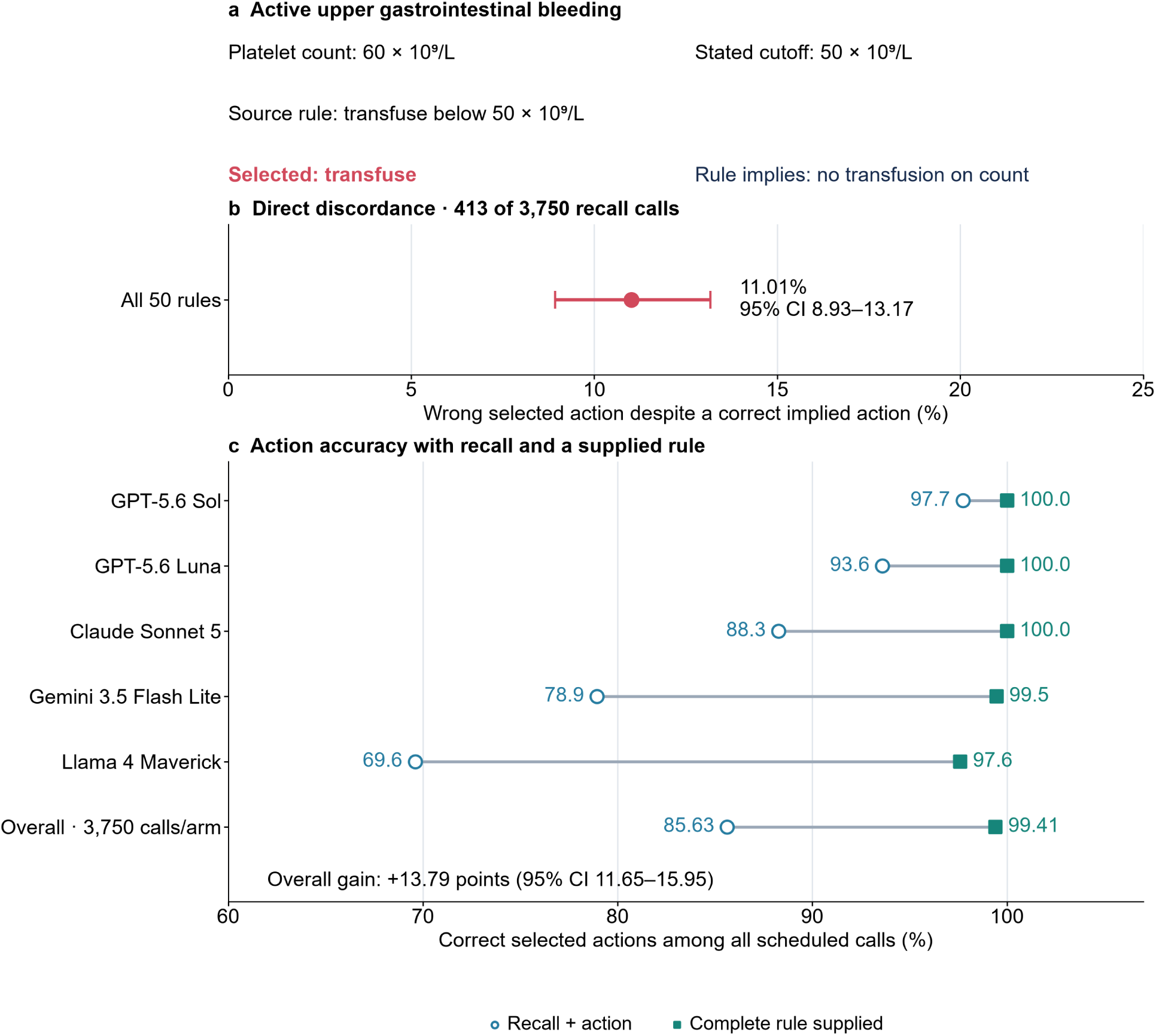
| Models can select the wrong action despite stating a cutoff that implies the right one. **a**, Observed Claude Sonnet 5 response for active upper gastrointestinal bleeding at a platelet count of 60 × 10⁹/L. The model stated 50 × 10⁹/L but selected transfusion; the source comparator is strictly below the cutoff. **b,** Strict discordance occurred in 413 of 3,750 scheduled recall-arm calls across 50 rules (11.01%; 95% CI, 8.93–13.17). A stated cutoff need not exactly equal the published cutoff to imply the correct action at a given case value. **c,** Action accuracy for the five fixed models and overall; each model contributes 750 calls per condition. Open circles denote recall plus action, filled squares the complete supplied rule. Model order is fixed. The overall paired accuracy gain is 13.79 percentage points (95% CI, 11.65–15.95), including 463 valid wrong-to-right changes and 54 invalid-to-correct recoveries. Intervals use 100,000 whole-rule bootstrap resamples and describe rule variation conditional on these models. Source values and the planned subset analysis are in Tables S67–S71.

Across the 50-rule experiment, this strict discordance occurred in 413 of 3,750 recall-arm calls (11.01%; 95% CI, 8.93 to 13.17), affecting 44 rules. Supplying the complete rule raised accuracy from 3,211 of 3,750 calls (85.63%) to 3,728 of 3,750 (99.41%): an improvement of 13.79 percentage points (95% CI, 11.65 to 15.95).

Among valid pairs, 463 changed from wrong to right and none from right to wrong. Another 54 invalid recall responses became correct supplied-rule responses, accounting for the remaining accuracy gain. Discordance varied across the five models, from 0.93% to 26.00% (Supplementary Table S68). Supplying the complete rule repaired most errors in this experiment, including errors that could not be attributed simply to an incorrect stated cutoff.

The earlier rule experiment showed that recalled cutoffs could also move with the patient value: 32.4% of recall case sets and 37.9% of recall-plus-calculation sets varied across the five values. Inferred actions reversed in the wrong direction in 7.6% and 11.8% of sequences, versus 0.2% with the cutoff supplied (Supplementary Figure 25). Its prespecified near-versus-far comparison found 8.47% versus 6.56% critical action errors (difference, 1.91 percentage points; 95% CI, 0.40 to 3.55; P=0.011; Table 1). Direct selection therefore revealed errors beyond those inferred from a reported number.

### Visible caution did not consistently identify numerical errors

The answer-or-decline instruction yielded valid numbers for 59.4% of requests, accuracy of 32.9% and very-large errors for 15.7%. Best-estimate increased these to 86.6%, 45.2% and 24.9%. Among requests answered under both instructions, accuracy changed little. Very-large errors recurred across all three repetitions in 21.2% of complete groups and retained their direction in 99.1%. Refusal, stated ranges and verification therefore did not consistently expose the failures (Supplementary Figures 18, 20–22).

## Discussion

Language models often reflected the numerical structure of clinical evidence yet failed to produce the answer required by a changed population or patient. They also selected incorrect actions when their stated cutoffs implied the correct actions. Numerical knowledge, adaptation to the case and action selection each require evaluation.

These errors can be difficult to notice. A probability may fall within the published literature’s range while being wrong for the population described. In the patient-level AUDIT-C example, movement from 0.69 to 0.47 showed sensitivity to the changed context, but calculation required 0.401. A decision threshold between those values would yield different choices.[35] The action experiment demonstrates the additional need to check the choice itself, even when the reported cutoff appears correct.

Quantitative clinical relationships complement evaluations based on expert judgment and conversational quality.[7,9,14,36] A prevalence change predicts a change in predictive value; patient information specifies a different target; an authoritative cutoff should remain fixed. The selected action can then be checked against the cutoff and comparator. Each relationship supplies a verifiable outcome without requiring a language-model judge.

The repairs identify practical development targets. Explicit inputs and calculation requests improved population and patient estimates, and supplying the complete rule raised direct-action accuracy to 99.41%. Clinical systems could make evidence selection, numerical computation and rule application explicit and verify their agreement before presenting an answer. The supplied-rule prompt combined the cutoff, comparator and action map, so its effect does not isolate which component produced the improvement. Neither prompting intervention was a deployed retrieval or calculation system.

Clinicians also struggle with conditional probabilities.[37,38] Here, the same models performed better when the operation was specified than when it had to be inferred from clinical wording. Stronger performance on matched nonmedical arithmetic suggests that clinical framing contributes to the difficulty without establishing a uniquely medical mechanism. Studies of public use and human– model interaction address a further question: how these responses influence users.[39,40,41]

These were controlled, single-turn text experiments without tools or patient outcomes. The evidence corpus was uneven, empirical patient targets were subgroup prevalences, and model panels differed across experiments. Rates may change with future models and clinical workflows.

Clinical AI intended to act should demonstrate the entire sequence: identify the applicable evidence, compute the case-specific quantity and select the action dictated by the governing rule. NUMBERS provides separate tests of those steps and shows that explicit numerical and rule-based inputs can substantially improve them.

## Data and code availability

Source datasets are identified in the references. The Supplementary Appendix and numerical workbook provide the methods, model configurations and source values for the reported results. The accompanying Supplementary Data and Code archive contains parsed benchmark and patient-level responses, direct-action records, frozen protocols and analysis code. Its contents and file check-sums are described in the archive README and manifest. Raw provider transcripts are excluded because they can contain private research or account information; the shared response files retain the numerical values, response classifications and identifiers needed to link them to the documented scoring rules.

## Author information

### Author contributions

Conceptualization: S.H., J.W.J., J.N.G. and A.S.R.; Methodology: S.H., J.W.J., P.S., S.L., J.N.G. and A.S.R.; Software: S.H. and P.S.; Validation: S.H., J.W.J., P.S., A.G., P.J.S., A.M., S.L. and J.N.G.; Formal analysis: S.H. and P.S.; Investigation: S.H., J.W.J., P.S., A.G., P.J.S., A.M. and S.L.; Data curation: S.H., P.S., A.G., P.J.S. and A.M.; Visualization: S.H. and P.S.; Writing, original draft: S.H.; Writing, review and editing: all authors; Supervision: S.H., J.N.G. and A.S.R.; Project administration: S.H.; Funding acquisition: S.H.

### Correspondence

Correspondence should be addressed to Shuhan He.

### Use of AI-assisted tools

OpenAI Codex and Anthropic Claude Code assisted with analysis code, code-generated figures and manuscript drafting. The authors are responsible for the scientific content and the decision to submit. These tools are not authors.

## Data Availability

Source datasets are identified in the references. The Supplementary Appendix and numerical workbook provide the methods, model configurations and source values for the reported results. The accompanying Supplementary Data and Code archive contains parsed benchmark and patient-level responses, direct-action records, frozen protocols and analysis code. Its contents and file checksums are described in the archive README and manifest. Raw provider transcripts are excluded because they can contain private research or account information; the shared response files retain the numerical values, response classifications and identifiers needed to link them to the documented scoring rules.

