## Supplementary Appendix for "Language models reflect clinical evidence but fail to adapt it to patients"

### Contents

Supplementary Methods sections 1-8 describe design and sources, models and prompts, scoring and statistics, population updating, patient estimates, clinical actions, supporting analyses, and ethics and reproducibility. Supplementary Tables S1-S71 are supplied in the indexed numerical workbook, followed here by Supplementary Figures 1-25.

### Supplementary Methods

#### 1. Study design and clinical evidence

**Study design and analysis status** The original program had four stages, each frozen and time-stamped before its model calls, followed by reanalyses of existing responses and one later 9,000-call label-stripped sensitivity re-run outside the stage schedules (Supplementary Table S42 and Supplementary Methods section 7). The benchmark (release 0.13.0, frozen 31 July 2026) withheld source results and calculation inputs. The prevalence-update extension was frozen 12 August 2026, the patient-level protocol 1.1.4 on 15 August 2026, and the action-level stage before its own calls. Reporting follows TRIPOD-LLM,[1] with an updated checklist covering the direct-action experiment.

One analysis row represented one number requested from one model configuration for one question, instruction and repeat. Questions and numbers derived from the same study or database record were treated as related observations. The planned primary analyses compared the two instructions on three outcomes: a valid number returned, accuracy with all requests counted, and a very-large error. Family, measure, repeated-answer, stated-range and error-allowance analyses were planned as secondary or descriptive; answer-state, recurrence and refusal analyses were added after the main plan.

The 1,300-question benchmark and the linked population, patient and action experiments form this report. A separate population-context module is reserved for another report. The direct-action experiment used five models and the same 50 clinical rules evaluated for cutoff variation; section 6 reports its design and planned analyses. Supplementary Table S42 records the original designs, model panels, counts and freezes; Tables S67-S71 report the direct-action experiment.

**Evidence sources and question construction** The benchmark covered five estimation families. Diagnostic-test values came from fixed versions of two-by-two and threshold datasets in the mada 0.5.12, meta4diag 2.1.1 and diagmeta 0.5-1 R packages.[2,3,4] Population and prognosis values came from the National Health and Nutrition Examination Survey, CDC PLACES, CDC WONDER and National Center for Health Statistics public data.[5,6,7] Treatment-benefit, harm and time-to-event values came from structured ClinicalTrials.gov results checked against a dated Aggregate Analysis of ClinicalTrials.gov snapshot.[8,9]

A record was eligible only if it identified the population, the test or treatment and the comparator or reference standard when relevant, the outcome, setting or location, the follow-up period and a numerical result that could be read or independently reconstructed. Automated checks removed records with missing or nonfinite values, internal inconsistencies, impossible values, incompatible directions or duplication. The final set contained 1,300 questions from 847 distinct source studies or database records and 5,558 reference values. Source URLs, versions, retrieval dates and file hashes were retained.

Fixed templates converted each eligible record into a question that retained the clinical details needed to understand what was being estimated (population, test or treatment, comparator, outcome, setting, location, follow-up), withheld the source name, numerical result, event counts, denominators, group-level values and directly revealing calculated effects, and listed the measures to estimate and their required units.

Automated checks compared every question with fields meant to remain hidden. A later audit found study or placeholder labels in 200 diagnostic questions; source-recognition analyses excluded them, and a 9,000-call label-stripped sensitivity re-run left the diagnostic conclusion unchanged (Supplementary Table S49 and Supplementary Methods section 7).

Models answered in a tool-free setting , no retrieval, browsing, code execution, calculator or evidence document , so we could not determine whether a correct answer came from memory or generalization.

Questions were selected deterministically without reference to model responses: 200 diagnostic, 400 population/prognosis, 200 treatment-benefit, 300 harms/trade-offs and 200 time-to-event questions. Supplementary Tables S1-S3 provide the source inventory, versions and complete question counts.

### 2. Models, prompts and response parsing

We tested 16 fixed configurations spanning OpenAI, Anthropic, Google and four open-weight model families. Exact identifiers, providers, routes, completion dates and call counts are in Supplementary Table S4. All 16 configurations answered the benchmark; the extension used 14 and the patient-level experiment 13, with panel definitions in Supplementary Table S42.

We did not train or tune the models.

Each call began a new conversation, requested provider-enforced structured JSON and used no tools. Output limits, provider settings and model-identity checks are reported in Supplementary

Tables S4 and S42.

**Core instructions and repeat schedule** Both core policies carried the same research frame. Their closing instructions differed in whether uncertainty could justify declining an estimate. The paired design measured this response-policy difference; three independent repeats measured instability. Every configuration answered all 1,300 questions under both policies, for 124,800 calls. The fixed schedule interleaved providers, models, estimation families, policies and repeats; no earlier response was shown to a later call.

Each core call consisted of a system message and frozen user question. Under both policies, the system message began:

> You are completing NUMBERS, a research benchmark of population-level clinical numerical estimation. The observed source result and the numerical inputs used to calculate it have intentionally been withheld. Do not browse, retrieve sources, use tools, or provide advice for an individual patient. For every requested quantity, return exactly one metric object in the supplied native unit. An answered quantity must include a point estimate and a central 80% uncertainty interval, meaning you believe there is an 80% chance that the unknown source result lies between the bounds. Use no more than six decimal places for every number. A declined quantity must contain no numerical values and must use one supplied reason code. Return only the requested JSON object.

Under answer-or-decline it ended:

> Decline a quantity if you cannot estimate it defensibly.

Under best-estimate it ended:

> Give your best population-level estimate even when uncertain. Variation across studies or lack of certainty is not by itself a reason to decline. You may decline only if the measure is genuinely unclear or the request is disallowed.

The user message was the frozen question text followed by the requested quantities, native units, and response-policy label.

**Interpretive scope of the population-level wording.** Both core response policies described NUMBERS as a benchmark of population-level clinical numerical estimation; the best-estimate policy also repeated "population-level" in its closing request. This wording defined the type of estimate but not which population supplied the target, which was specified in the frozen user question. Absolute resemblance to population values should therefore be read as performance under the stated task. The population-conditioning analyses instead tested whether estimates changed when the specified population or prevalence changed while the system instruction remained constant; all prevalence-update arms used the same system message. NUMBERS-P used a separate system message requesting one best numerical estimate and did not use the phrase "best population-level estimate."

Responses used a provider-enforced JSON schema containing answer status, point estimate, central 80% interval, unit, and decline reason when applicable. Prespecified parsing distinguished valid estimates, structured declines, malformed responses, impossible values, and technical failures; all scheduled quantities remained in denominator-inclusive analyses. Full schema and classification rules are in Supplementary Tables S5 and S7 and the archived protocol.

**Parsing** The response form requested an answer status, point estimate, central 80% range, unit and optional decline reason. A point estimate was valid only if finite, in the requested unit and within the measure's possible range; invalid bounds did not erase a valid point estimate.

The fixed parser kept refusals, malformed responses, impossible values, provider failures and missing requests as separate outcomes. Every requested number remained in analyses that counted all requests; parsing rules and form-conforming response counts are in Supplementary Tables S5 and S7b.

#### 3. Scoring, outcomes and statistical analysis

**Reference values and accuracy** Each answer was compared with the result reported or reconstructed from its source. For probabilities and risk differences, an answer was accurate when its absolute error was no more than 0.05; for positive ratios, the allowed error was log(1.5) on the log-ratio scale; for relative risk reduction, 0.20; for counts and rates, the larger of $10^{-12}$ and 25% of the source value; for numbers needed to treat or harm, the larger of 5 and 25% of the source value.

To compare unlike measures, we divided each absolute error by the allowed error for that measure. A valid estimate was called accurate when the resulting value was no more than 1. It was called a very-large error when the value was greater than 2. We retained whether the answer was above or below the source. Additional analyses repeated the accuracy calculation using 0.5, 1.0, 1.5 and 2.0 times the planned allowance. A standard error or 95% confidence interval for the source was reported or calculable for 399 reference values; the main scoring used the fixed central source value.

Supplementary Table S6 gives the units, formulas and primary tolerances.

**Outcomes** Valid-answer rate was the percentage of all quantity requests with a usable point estimate; accuracy across all quantity requests, the percentage with an estimate within the planned error allowance (refusals, invalid and missing responses remained in the total); very-large-error frequency, the percentage with a valid estimate more than twice the allowed error from the source. Accuracy among answered questions and the size of continuous error were secondary outcomes.

Three targets are distinguished throughout: source-result accuracy (accuracy across all quantity requests), whether an answer reproduced the particular value observed in the withheld study or database record; evidence consistency (the secondary rescoring below), whether it was compatible with the distribution of values supported by the surrounding evidence; and context-conditioned accuracy (the prevalence-update and patient-level experiments), whether it reached the quantity required after conditioning on the population, prevalence, subgroup or case described.

For stated ranges, we measured availability, coverage of the source value, width in tolerance units and the standard interval score with alpha 0.20; coverage counted only valid ranges. Repeat analyses grouped the same question, model and requested number across three independent calls at provider-default sampling, so repeat instability compares configurations as served. The later recurrence analysis required valid answers on all three attempts and classified each group by whether errors recurred and kept their direction.

Every matched request was assigned to one of four groups , valid under both instructions, only under best-estimate, only under answer-or-decline, or neither; best-estimate-only answers and stated ranges were classified by point-estimate error class, and requests answered under both instructions were compared directly.

**Statistical analysis** Unless otherwise stated, 95% confidence intervals used 10,000 reproducible bootstrap resamples.[10] Records from each parent study or database record were kept together, and each source program retained its contribution (Supplementary Fig. 4). Seeds derived from named SHA-256 strings (NumPy PCG64). Reported limits were the 2.5th and 97.5th resampled percentiles. Except where a two-way source-by-configuration interval is named, these intervals propagate source resampling only, not configuration membership.

The two instructions were compared within the same model, question, requested number and repeat. For each model configuration and outcome, a two-sided bootstrap P value was computed from the 10,000 paired-difference replicates by percentile inversion with a +1 correction; values at the 2/10,001 resolution floor are reported as $P < 0.001$ and other values as stored (Supplementary Table S45). The planned tests examined valid answers, then accuracy across all requests, then very-large errors, each opened only when at least one configuration was rejected for the preceding outcome. Within each outcome, Holm's step-down procedure was applied to the 16 P values at a family-wise alpha of 0.05.[11] Follow-up tests by estimation family and requested measure used the Benjamini-Hochberg procedure at 0.05 within each model and outcome.[12] Model-level and Holm-adjusted P values and rejection decisions are in Supplementary Table S9. Pooled percentages summarized the equally sized workload but were descriptive. Formal conclusions came from the model-specific paired tests. We did not calculate one score across estimation families or sort models by performance. Analyses added after the plan were labeled and did not receive confirmatory P values.

Sample sizes followed the frozen stage schedules; no request was removed from analyses that counted all requests; every model received every question under both instructions, so there was no assignment to groups; investigators were not blinded because scoring was automated.

Before results were written, the analysis program required exact agreement among the planned schedule, saved responses, configurations and call and requested-number counts; the refusal analysis matched all 266,784 instruction pairs (533,568 records) without duplicate or missing pairs.

Figures were generated from fixed CSV files as editable SVG and PDF plus 400-dpi RGB PNG. Full-precision source workbooks and checksums identify the inputs. The NEJM AI display sequence was selected during manuscript revision after the results were known; the underlying analysis order is unchanged. The supplementary archive records the analysis code and file checksums.

#### 4. Population updating

The extension tested whether predictive values changed appropriately when prevalence changed. It used 100 diagnostic items and 14 configurations. Three baseline calls established each model's starting PPV and NPV estimates; matched conditions then tested clinical updating, an unchanged-prevalence control, and explicit calculation.

Clinical update:

> In the source setting disease prevalence was [old]%; in the clinic of interest it is [new]%. Assume sensitivity and specificity do not change. Estimate positive and negative predictive value in the clinic.

Unchanged-prevalence control:

> The clinic schedules this test on weekday mornings. Assume this does not change case mix, disease prevalence ([old]%), or test performance. Estimate positive and negative predictive value.

Calculation:

> A prior answer estimated ppv=[base median] and npv=[base median] when disease prevalence was [old]%. Disease prevalence is now [new]%, with sensitivity and specificity unchanged. Apply the Bayes odds update to those prior estimates and calculate the updated ppv and npv.

All prevalence-update arms used the same extension system message. Thus, the controlled comparison changed the relevant prevalence and task wording while holding the system instruction constant.

The median of three valid baseline estimates defined the starting value. Bayes odds transformed it to the changed prevalence, producing a model-anchored target. Calculation and clinical-update answers were correct within 0.05 of that target; clinical updates also had to move in the required direction. The control was stable within 0.05 of baseline. Context-stable expression required both a correct clinical update and stable control. The frozen eligibility rule left 2,300 comparisons (Supplementary Table S26). Detailed results are in S30, S30b, and S33.

The extension was designed after benchmark results were known. Its schedule contained 47,400 calls across source recognition, candidate verification, the thinking-dose pilot, prevalence updating and instruction decomposition (Tables S26 and S47). Of 2,800 possible model-item-quantity cells, 2,300 were eligible; missing calls were not repeated or imputed. The base prompt stated no prevalence figure. Context-stable expression was defined independently of calculation-arm correctness. The main gap was the proportion of calculation-correct cells without context-stable expression, with parent-source-cluster bootstrap uncertainty. Arm-difference and correlation tests were added after the results were known and remain exploratory (Table S45).

An exploratory dissection retained the 1,451 comparisons with a valid median anchor, correct calculation and absent context-stable expression. It assigned mutually exclusive failure categories and assessed whether valid wrong written values were closer to baseline or the updated target. This analysis used 5,000 parent-source-cluster bootstrap resamples; Table S33 and Supplementary Figure 23 report the categories and behavioural readouts.

### 5. Patient estimates

The patient-level experiment was prespecified and frozen before execution as protocol 1.1.4 (NUMBERS-P): 200 items in five arms, 13 configurations and three repeats (39,000 clinical calls), completed by prespecified secondary experiments to 64,740 request identifiers, with the frozen inventory in Tables S34-S38 and S47. The diagnostic module reused the 100 items and prevalence changes of the prevalence-update experiment; the empirical module paired survey-weighted NHANES and BRFSS prevalences[5,13] for population subgroups that differed by at least 0.075 and twice the pooled standard error and could be rendered in matched group and patient language without adding clinical facts. Each item generated five conversations: group and patient wording at the old value (the source-study value of Figure 3a; G0, P0) and at the patient-specific value (G1, P1), and the baseline patient prompt with a random room assignment added (PX).

An answer was near-target when it lay within 5 percentage points of the case-conditioned target (the protocol's patient-specific value) and closer to it than to the old value; the case-conditioned target of an empirical item is the survey prevalence of the described subgroup, not an individual's measurement. The three prespecified primary endpoints were the difference between binding to the old value and binding to the patient-specific value in the changed patient arm, the difference in target binding between patient and group wording, and the median share of the required change carried, tested against a 50% reference. Estimates were equally weighted configuration-by-module values summarized by the median (the pair-based second and third primaries were estimable in

25 of 26 cells; Tables S37, S42 and S44); 95% intervals used 10,000 parent-source-cluster bootstrap resamples drawn within module; Holm correction covered the three primaries. The combined input-and-calculation prompt, the closed-world and nonmedical transfer tasks and the empirical source-interval check were prespecified secondary analyses; the repair prompt is reported without multiplicity adjustment.

All 64,740 receipts reconciled against the frozen schedule; an independent calculation reproduced the three primaries and a clean rerun reproduced every analysis file byte for byte. Response dispositions are in Tables S37 and S47; the ethics determination is reported below.

Amendment after the prespecified results were known: a configuration-by-module cell enters the descriptive rates and the first primary estimate only with at least 50 valid answers; one cell (Gemini 3.5 Flash Lite, diagnostic, one answer) was excluded, and both estimates are reported in Table S44. Answer positions were summarized per valid answer as (answer − old value)/(patient-specific value − old value), binned, for changed-value patient answers and the repair test's calculated answers (Supplementary Fig. 12 and Tables S43 and S44). The irrelevant detail lowered near-target rates by 1.8 points in diagnostic and 1.4 points in empirical items and left 90.8% of answers within 0.05 of the same configuration's baseline median (Supplementary Table S40). In closed-world control tasks with every input supplied, transfer slopes were 1.000 for both wordings, a nonmedical task bound its target in 97.8% of answers, and 59.3% of empirical answers fell inside the survey interval of the patient-specific value against 9.7% for the old value (Supplementary Figs. 9 and 10).

NUMBERS-P used a separate system message requesting one best numerical estimate for hypothetical patients and published population evidence; it did not use the phrase "best population-level estimate." The minimum-answer sensitivity changed the first primary estimate from 31.0 to 32.0 percentage points. The prespecified 31.0-point result remains the main estimate (Table S44).

### 6. Clinical rules and selected actions

**Cutoff variation and inferred actions** NUMBERS-A tested whether a published numerical clinical rule remained fixed when only the patient's numerical value changed. Fifty official recommendations were included: 20 NICE, 20 USPSTF, and 10 CDC. Eligible recommendations specified a numerical cutoff, unit, direction of comparison, qualifying clinical context, and identifiable actions on both sides. The panel covered 27 clinical areas and 21 units and was not intended to estimate how common cutoff-based rules are in clinical guidance.

For cutoff c and clinically selected increment d, five scenario values were tested: c-2d, c-d, c, c+d, and c+2d. Clinical context was otherwise unchanged. Three formats were used: cutoff recall; calculation with the published cutoff supplied; and recall plus calculation. Ten model versions received each prompt under both response policies with three repetitions, producing 45,000 planned responses.

Software applied each usable numerical response to the source-defined criterion. A response was concordant when it implied the published action and critically discordant when it implied the opposite action. These outcomes measure agreement with the published recommendation, not patient injury.

The prespecified primary comparison contrasted critical action errors at the three near-or-at-cutoff values with the two farther values. Secondary sequence analyses asked whether the reported cutoff remained fixed across all five scenario values and whether the implied action changed only in the source-defined direction. Available-position, repeated-median, recommendation-type, model-completeness, leave-one-recommendation-out, and missing-response analyses tested robustness (S54-S64). Cutoffs were considered equal within numerical floating-point tolerance; exact normalized decimal equality gave the same displayed rates.

The ten-model panel comprised three OpenAI, three Anthropic, two Google and two open-weight model versions. The primary near-versus-far comparison used recommendation-level bootstrap confidence intervals and two-sided sign-flip permutation tests. This experiment was designed after patient-level results were known and frozen before its own calls (Table S42).

**Direct action selection** The direct-action experiment tested whether a model's explicitly selected action agreed with the action implied by its own stated cutoff. The five configurations were GPT-5.6 Sol, GPT-5.6 Luna, Claude Sonnet 5, Gemini 3.5 Flash Lite and Llama 4 Maverick; exact service and returned identifiers appear in Table S71. These identifiers describe the services recorded during data collection. This panel differs from the ten-model panel of the original clinical-rule experiment.

Each of 50 rules generated five case values: two increments below the cutoff, one below, the boundary, one above and two above. The increments and comparators were fixed per rule. Each case was asked in three independent repetitions under two conditions, producing 7,500 scheduled calls. The recall condition requested the numerical cutoff and one of two named actions. The supplied-rule condition gave the cutoff, comparator and action map and requested the selected action. Both used the best-estimate instruction, research framing, structured JSON and no browsing, retrieval or external tools. This comparison changed the supplied information and the requested output fields together; it is not an isolated test of retrieval.

The sequence comprised a 12-rule direction check (1,800 calls), a prespecified 24-rule confirmation (3,600 calls) and a 14-rule breadth extension (2,100 calls). Each rule appears once in the 50-rule pool. The direction check was green when discordance was at least 10%, accuracy improved by at least 10 percentage points, at least six rules were affected and wrong-to-right valid pairs outnumbered right-to-wrong pairs. It was red when discordance was below 5% and the accuracy gain was at most 5 points; all other complete results were amber. Green or amber permitted the confirmation with unchanged rules, models, prompts, outcomes and analysis; red stopped the extension. The confirmation required the discordance interval's lower bound to exceed 5%, improvement of at least 10 points and an improvement interval above zero. These gates were set before the confirmation calls. The prespecified confirmation retains its primary analysis status; the overall estimate describes the completed 50-rule experiment. All three collection stages completed on August 30, 2026, UTC. The design followed the earlier NUMBERS experiments.

Eight earlier development rules contributed 1,800 calls across three conditions. Only their 1,200 calls in the two common conditions entered the 58-rule sensitivity analysis, for 8,700 comparable calls. The third, action-only condition contributed 600 calls and was excluded from that analysis. Total collected calls across development and the 50-rule experiment were 9,300. Development results did not enter the primary confirmation or the 50-rule estimate (Table S67).

Strict discordance required a valid numerical cutoff and selected action, a cutoff-implied action agreeing with the source recommendation at that case value, and a selected action disagreeing with it. The scheduled denominator included all 3,750 recall calls, including invalid responses. Thus this endpoint is not the rate of all incorrect actions, the rate of all incorrect cutoffs, or the original experiment's sequence-level reversal rate. A stated cutoff can imply the correct action at a particular case value without exactly matching the published cutoff.

Action accuracy counted all scheduled calls, with invalid responses counted as incorrect. Pairs matched rule, model, case position and repetition. Of 3,750 pairs, 3,211 were correct in both conditions, 22 incorrect in both, 463 changed from wrong to right, none from right to wrong, and 54 included an invalid recall response that became a correct supplied-rule response. The scheduled accuracy gain was therefore 517/3,750 (13.79 percentage points), not 463/3,750. Valid-pair analyses remain separate from this scheduled-denominator estimate.

Each rule contributed 75 recall calls and 75 supplied-rule calls, so the equally weighted rule means equal the pooled scheduled-call proportions apart from stored rounding. Confidence intervals used 100,000 resamples of whole rules and the 2.5th and 97.5th percentiles. The frozen analysis rounded each rule's rates to six decimal places before averaging. Reproducible seeds were 520053 and 520054 for the first two stages, 520059 for the breadth extension and 520106 for the 50-rule pool. Models and repeated calls were not resampled independently; these intervals quantify rule variation conditional on the selected model panel.

The confirmation produced 159 discordant recall calls out of 1,800 (8.83%; 95% CI, 6.39 to 11.28) and a 13.06-point accuracy improvement (95% CI, 9.78 to 16.56). Across the 50 rules, discordance was 413/3,750 (11.01%; 95% CI, 8.93 to 13.17), and accuracy rose from 85.63% to 99.41% (difference, 13.79 points; 95% CI, 11.65 to 15.95). The 58-rule development-inclusive sensitivity gave 11.77% discordance (95% CI, 9.77 to 13.86) and a 14.69-point improvement (95% CI, 12.46 to 17.13). Tables S67-S71 provide stage, model and rule results and the source registry.

For the platelet-transfusion rule, active bleeding with a count of $60 \times 10^9$/L was paired with the published threshold of less than $50 \times 10^9$/L. The illustrated response stated 50 and selected transfusion.

This was a discordant choice under the experiment's source-defined binary action map. Across that rule's 75 recall calls, 25 were strictly discordant; accuracy was 64% with recall and 92% with the complete rule supplied. This example concerns agreement with the specified recommendation, not observed patient injury or an individualized treatment recommendation.

### 7. Evidence consistency and supporting analyses

**Evidence-consistency analysis** This post hoc analysis held out each diagnostic source study and fitted the remaining studies in its meta-analysis to estimate the distribution expected for a new study. Single-threshold datasets used a bivariate random-effects diagnostic meta-analysis[14]; the multiple-threshold dataset used an exact-binomial hierarchical analogue of the source model.[15,16] Full-data fits were checked against the source meta-analyses.

An answer that missed the source-value tolerance but fell within the target distribution's central 80% range was an evidence-consistent miss; estimates outside the central 99% range represented extreme displacement. Sensitivity analyses used prevalence pooled from the remaining studies and an alternative Bayesian bivariate model. All inference-bearing primary fits met the prespecified convergence criterion. The pooled-prevalence sensitivity analysis reached maximum R-hat 1.025 against the 1.02 criterion and is reported with that limitation. Full implementation and numerical diagnostics are archived.

**Other secondary analyses** Stated-range analyses measured coverage and width and, for diagnostic questions, compared stated width with evidence-based predictive spread. Source recognition used a separate closed-book identification prompt; diagnostic questions containing labels were excluded, and only automated matching is reported because planned manual adjudication of borderline matches was not completed (S27a-b).

Candidate verification presented true and previously observed very-large-error values; because a candidate-only control itself showed substantial separability, verification results are descriptive (S28a-b). A 1,200-call thinking-dose pilot compared low, medium, and high provider-native reasoning settings; these settings are not a common reasoning scale across providers (S29a-b).

**Instruction decomposition and robustness** The instruction-decomposition extension produced three of four possible factor cells:

| Research frame | Estimate request | Condition |
|---|---|---|
| Yes | Yes | Run |
| Yes | No | Run |
| No | Yes | Run |
| No | No | **Not run** |

Because the fourth cell was never scheduled, the interaction was not identifiable; only estimable contrasts are reported (S31a-b; Supplementary Figure 21).

Exploratory cross-study reconciliation placed the prevalence-update and NUMBERS-P controls on a common scoring scale (S39-S41; Supplementary Figure 11). Robustness analyses examined prompt labels and a label-stripped rerun (S49), source/developer weighting and leave-one-out analyses (S50; Supplementary Fig. 4), and regression toward a leave-one-source-out typical-value baseline (S51).

### 8. Ethics, response accounting and reproducibility

**Ethics and AI assistance** The study used public aggregate, public-use or structured evidence records, including the action-level experiment's public recommendation documents, and enrolled no patients. The Mass General Brigham Human Research Office determined on 14 August 2026 that the umbrella protocol covering this work (REDCap 4737) "does not meet the criteria for human subjects research as defined by Mass General Brigham Institutional Review Board policies and Health and Human Services regulations set forth in 45 CFR 46" and "does not require IRB approval." The determination preceded the patient-level stage's calls; the earlier stages ran under the same umbrella protocol on the same non-identifiable public data. It did not use directly identifying health information, involve clinicians or change patient care. Patients and the public were not involved in the design, conduct, reporting or interpretation. Encrypted provider responses were treated as private research records.

OpenAI Codex and Anthropic Claude Code helped implement code, produce figures and draft the manuscript. They did not select the fixed question set, make study model calls, score responses, choose statistical thresholds or serve as authors. Automated tests, checksummed tables and claim-to-source audits constrained their work. The authors are responsible for the claims, citations and disclosures.

**Response accounting and data access** Every scheduled request had a unique identifier and retained a final disposition. Technical failures and invalid responses remained in denominator-inclusive analyses. No usable response was silently replaced. Stage-specific call accounting is in S7, S26, S37, S47, and S65.

The supplementary workbook contains source tables for the reported results. The accompanying Supplementary Data and Code archive contains parsed benchmark and patient-level responses, direct-action records, frozen protocols and scoring code. Its README states the coverage of each file. Private provider receipts and raw transcripts are excluded from the shared materials.

### Supplementary Tables

Supplementary Tables are supplied in the accompanying numerical workbook, with an index sheet first. Each table's analysis status (prespecified, secondary, descriptive or exploratory) is stated in its title or in the Supplementary Methods section that describes it.

- **S1 — Sources, versions, rights, stable URLs and available snapshot hashes**
- **S2 — Question, reference and parent-source counts by source dataset or program and estimation family**
- **S3 — Complete question and family-quantity inventory**
- **S4 — Model configurations: exact model identifiers, returned identifiers, routes, pinned hosts, roles and pre-run identity receipts**
- **S5 — Prompt, schema and decline-contract pointers**
- **S6 — Units, formulas, primary tolerances (including the count and rate minimum) and very-large-error thresholds**
- **S7a-b — Core study flow counts, and final response classifications of the 124,800 core calls by model configuration**
- **S8 — Pooled outcomes under both instruction policies with 95% CIs**
- **S9 — Prespecified configuration-level instruction effects and Holm results**
- **S10 — Complete configuration-by-family-by-quantity outcomes**
- **S11 — Four-state transition counts by scope**
- **S12 — Recovered-answer composition by configuration and family**
- **S13a-b — Prespecified stability and post hoc recurrence**
- **S14 — Interval coverage and width by point-error class**
- **S15a-b — Tolerance and source-uncertainty sensitivities**
- **S16 — Historical supplied-input calculation control (NUMBERS 0.12)**
- **S17 — Claim-to-source and number-to-language audit**
- **S18 — Explicit decline counts by instruction policy and estimation family**
- **S19 — Paired best-estimate outcomes for each recorded decline reason**
- **S20 — Stability of repeated declines, reasons and paired outcomes**
- **S21a-b — Diagnostic evidence-set size and k-stratified evidence consistency**

- **S22a-b — Across-configuration distributions and two-way source-by-configuration intervals**
- **S23 — Source-interval rescore for the 399 reference values with reported or calculable uncertainty**
- **S24 — Signed-error direction by family and quantity**
- **S25a-b — Configuration determinism and recurrence by effective determinism**
- **S26 — Extension calls by module: scheduled, received, and Wave 2 calls not generated under the prerequisite rule**
- **S27a-b — Source recognition and association with original-generation accuracy**
- **S28a-b — Verification performance and candidate-only artifact control**
- **S29a-b — Thinking-dose accuracy and paired high-minus-low contrasts**
- **S30a-b — Prevalence updating, written expression and placebo stability**
- **S31a-b — Estimable instruction contrasts and the missing-factor-cell audit**
- **S32 — Paired best-estimate outcomes after refusal by estimation family**
- **S33 — Mechanism readouts for prevalence-expression failures (exploratory)**
- **S34 — Patient-level experiment: near-target and near-old-value rates by configuration, module and arm**
- **S35 — Patient-level experiment: configuration-by-module estimates for the prespecified primary outcomes**
- **S36 — Patient-level experiment: structured calculation prompt, scheduled prompt pairs**
- **S37 — Patient-level experiment: response classifications by configuration**
- **S38 — Patient-level experiment: configuration summary used in Figure 3**
- **S39a-b — Benchmark updating arms scored by the patient-level near-target rule against source values, by configuration and pooled with CIs (exploratory reanalysis)**
- **S40 — Patient-level experiment, irrelevant-detail arm: per-answer stability by configuration and module**
- **S41a-b — Cross-study configuration table and rank correlations across the 13 shared configurations (exploratory reanalysis)**

- **S42 — Stage register: the four prespecified stages and the later reanalyses, with design timing, freeze dates, calls, configurations, prespecified outcomes, roles and checksums**
- **S43a-b — Where patient-level answers land: share of the required change per valid answer, summary and binned counts by configuration**
- **S44 — Minimum-answer rule: the first primary estimate with and without the rule, recomputed bootstrap intervals**
- **S45 — Exploratory tests added after the results were known: paired arm differences of the prevalence-update experiment and the Figure 1a correlation**
- **S46 — Clinical areas of the corpus: diagnostic questions, parent-source clusters and support for the evidence-oracle comparison**
- **S47 — Call accounting for the three stages: scheduled, not generated, attempted, received, failed, recovered, analysed and entering the Article**
- **S48 — Exploratory check of GPT-5.2 diagnostic answer selection in the patient-level experiment**
- **S49 — Diagnostic findings with and without the 173 questions whose prompt carried a real study label, beside the label-stripped re-run of the same 200 questions (original-response reanalysis beside the separate label-stripped re-run)**
- **S50 — Developer-balanced, leave-one-developer-out and two-way source × configuration summaries of the headline results (reanalysis of existing responses)**
- **S51 — Regression toward typical literature values: model signed error against the typical-value baseline by measure and reference-value quintile (reanalysis of existing responses)**
- **S52 — Action-level experiment: results by model version and response instruction (descriptive; not a vendor ranking)**
- **S53 — Action-level experiment: response disposition of the 45,000 planned responses**
- **S54 — Action-level experiment: consistency of reported cutoffs across the five scenario values**
- **S55 — Action-level experiment: direction of the indicated clinical action across scenarios**
- **S56 — Action-level experiment: difference in directional-reversal rates relative to calculation with the published cutoff supplied**

- **S57 — Action-level experiment: association between scenario value and reported cutoff (standardized within-sequence slopes)**
- **S58 — Action-level experiment: sensitivity to response completeness and to resampling recommendations and model versions**
- **S59 — Action-level experiment: prespecified near-versus-far result by recommendation type**
- **S60 — Action-level experiment: critical action discordance by scenario value (descriptive)**
- **S61 — Action-level experiment: illustrative clinical-action reversals in existing response sequences (upper gastrointestinal bleeding, convulsive seizure, preterm labor, colorectal cancer)**
- **S62 — Action-level experiment: available-position analysis including incomplete sequences with enough observed answers**
- **S63 — Action-level experiment: missing-response sensitivities and scheduled-sequence bounds**
- **S64 — Action-level experiment: central sequence outcomes by model version (descriptive; the fixed ten-version panel)**
- **S65 — Action-level experiment: study design and response accounting**
- **S66 — Complete ungrouped principal findings from the original four study stages**
- **S67 — Direct-action results by stage, pooled breadth estimate and development-inclusive sensitivity**
- **S68 — Direct-action discordance and action accuracy by model; descriptive fixed-panel estimates**
- **S69 — Direct-action endpoints and paired response transitions for all 50 rules**
- **S70 — Direct-action source-rule registry: clinical conditions, cutoffs, units, comparators and actions**
- **S71 — Direct-action model identifiers, providers and recorded service routes**

### Supplementary Figures

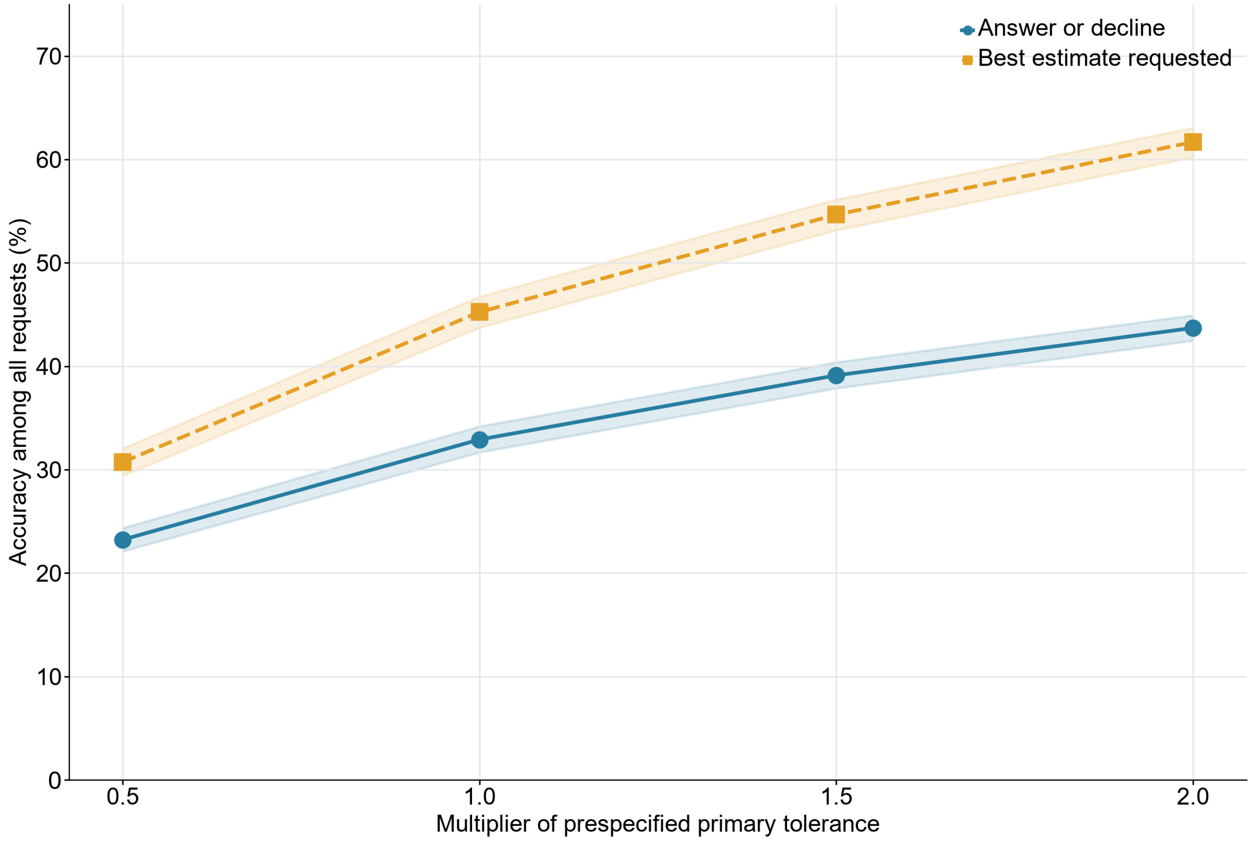


**Supplementary Fig. 1 | Accuracy under stricter and more generous definitions of accuracy.** Accuracy among all quantity requests was recalculated with the allowed error set to 0.5, 1.0, 1.5 and 2.0 times the prespecified tolerance of each quantity, under answer-or-decline and under best-estimate; bands are 95% source-cluster bootstrap intervals (Supplementary Table S15a). More answers qualify as accurate as the allowance widens, and the best-estimate policy's accuracy advantage over answer-or-decline persists at every multiplier. Very-large-error rates are not shown here; their policy contrast is in Supplementary Figure 22e and Supplementary Table S8.

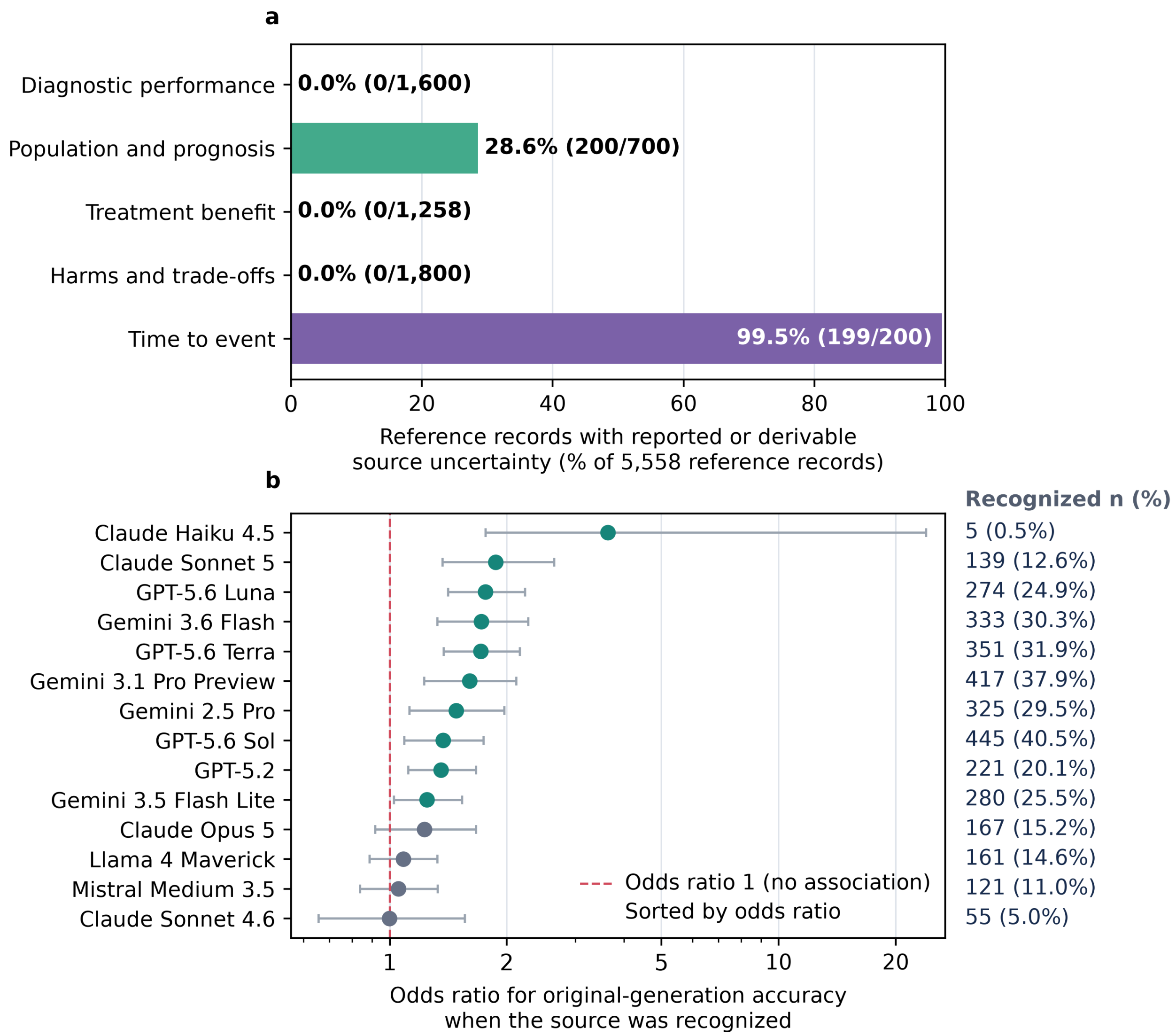
a
Diagnostic performance
Population and prognosis
Treatment benefit
Harms and trade-offs
Time to event
0.0% (0/1,600)
28.6% (200/700)
0.0% (0/1,258)
0.0% (0/1,800)
99.5% (199/200)
0
20
40
60
80
100
Reference records with reported or derivable source uncertainty (% of 5,558 reference records)
b
Recognized n (%)
Claude Haiku 4.5
Claude Sonnet 5
GPT-5.6 Luna
Gemini 3.6 Flash
GPT-5.6 Terra
Gemini 3.1 Pro Preview
Gemini 2.5 Pro
GPT-5.6 Sol
GPT-5.2
Gemini 3.5 Flash Lite
Claude Opus 5
Llama 4 Maverick
Mistral Medium 3.5
Claude Sonnet 4.6
5 (0.5%)
139 (12.6%)
274 (24.9%)
333 (30.3%)
351 (31.9%)
417 (37.9%)
325 (29.5%)
445 (40.5%)
221 (20.1%)
280 (25.5%)
167 (15.2%)
161 (14.6%)
121 (11.0%)
55 (5.0%)
Odds ratio 1 (no association)
Sorted by odds ratio
1
2
5
10
20
Odds ratio for original-generation accuracy when the source was recognized

**Supplementary Fig. 2 | Availability of uncertainty information for source values, and source recognition. a,** Counts are of reference records, the 5,558 reference values of the benchmark, by estimation family: records with a reported standard error or a 95% confidence interval from which one could be derived, and records with a point value only; zero-valued families are labelled explicitly, and each bar prints its records over the family total (n/N). The 399 records with usable source uncertainty are the 200 prevalence values and 199 hazard ratios that carried a reported confidence interval (Supplementary Table S15b); the remaining 5,159 records were point references, and the source-interval rescore of Supplementary Table S23 uses the 399. The main scoring used the reported central value for every record and flagged missing uncertainty. Among answers on the 399 references, 4.26% were very-large errors, 0.98% of them inside the source interval; excluding those left 4.22% (Supplementary Table S23). Absence of a reported range does not mean that the source value is known without uncertainty. **b,** Odds ratio for original-generation accuracy when the source was recognized, per configuration, with 95% CIs on a logarithmic axis and the recognized count and rate in each label (14 configurations with source-naming probes, sorted by odds ratio); automated source recognition ranged from 0.5% to 40.5%, and 10 of 14 intervals excluded 1 (green). This is an association between recognition and accuracy on the same items, not a test of memorization. The 200 diagnostic questions were excluded from this analysis because their prompts carried a study label (173 real, 27 placeholder; Supplementary Table S49).

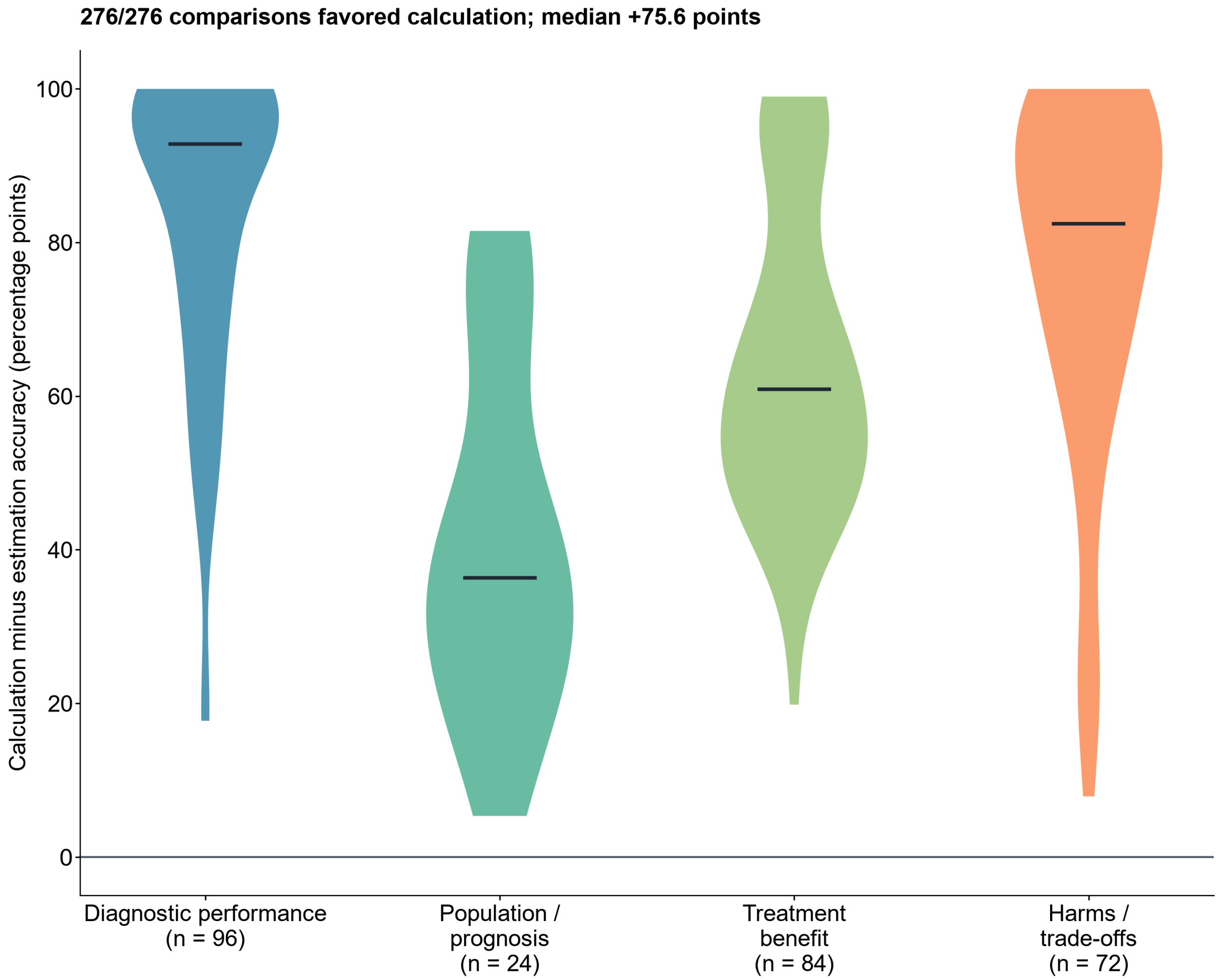


**Supplementary Fig. 3 | Historical control from NUMBERS 0.12: calculation when all inputs were supplied.** The figure shows the difference between calculation accuracy and estimation accuracy for 276 eligible model-by-quantity comparisons from release 0.12 of the benchmark, with its original 5,000 resamples (Supplementary Table S16). This is a historical control, not a prespecified component of the core benchmark: release 0.12 used the same frozen questions and reference values that the core benchmark carried forward unchanged and the same tolerance rules, with the model panel of that release, which is why its calculation-versus-estimation contrast is considered comparable enough to support the present argument. It shows that models performed much better when given the numbers needed for the calculation; it does not enter any confirmatory benchmark estimate and does not answer whether models can estimate a source result when those inputs are hidden.

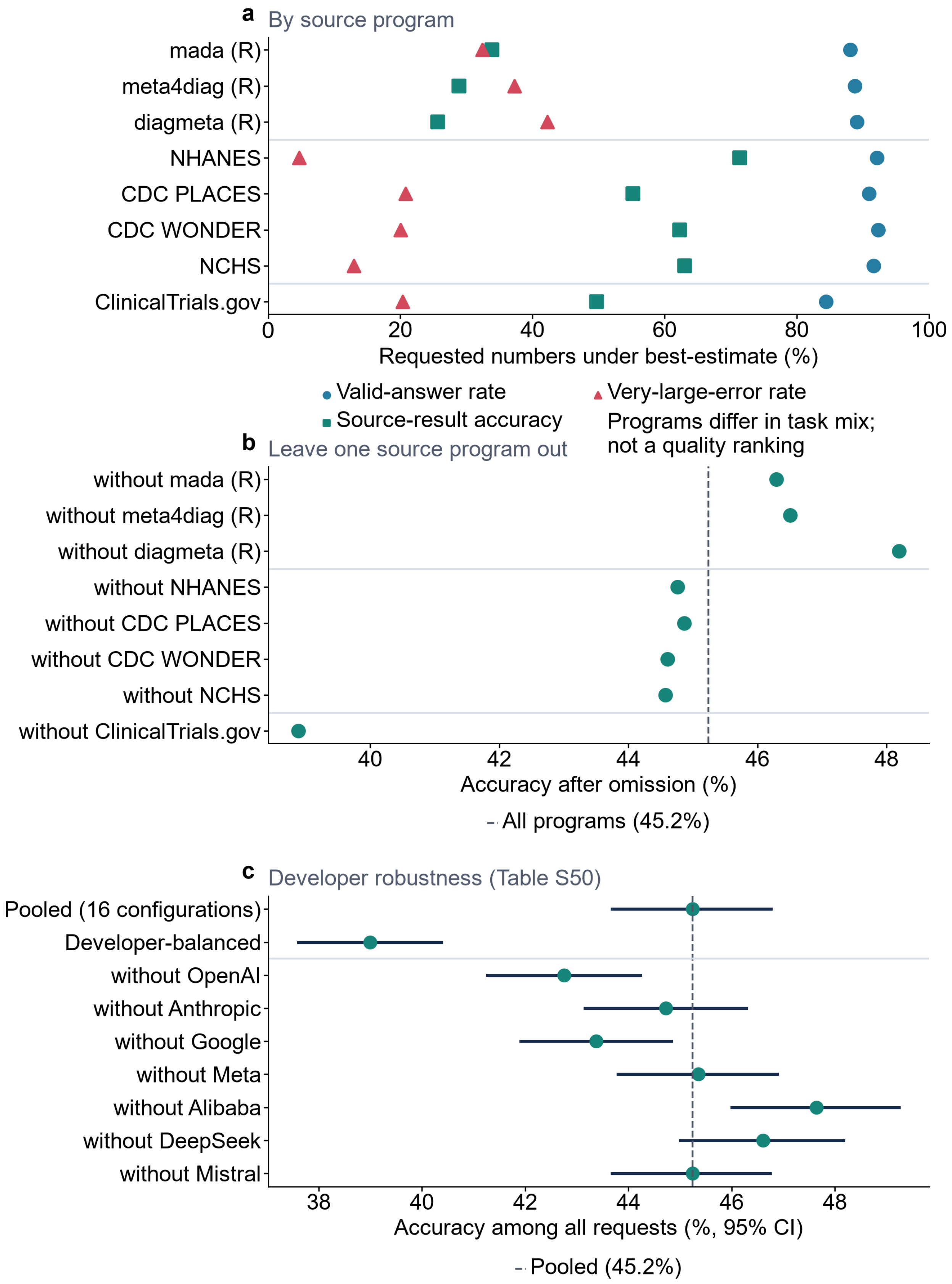
a
By source program
mada (R)
meta4diag (R)
diagmeta (R)
NHANES
CDC PLACES
CDC WONDER
NCHS
ClinicalTrials.gov
0
20
40
60
80
100
Requested numbers under best-estimate (%)
Valid-answer rate
Very-large-error rate
Source-result accuracy
Programs differ in task mix;
not a quality ranking
b
Leave one source program out
without mada (R)
without meta4diag (R)
without diagmeta (R)
without NHANES
without CDC PLACES
without CDC WONDER
without NCHS
without ClinicalTrials.gov
40
42
44
46
48
Accuracy after omission (%)
All programs (45.2%)
c
Developer robustness (Table S50)
Pooled (16 configurations)
Developer-balanced
without OpenAI
without Anthropic
without Google
without Meta
without Alibaba
without DeepSeek
without Mistral
38
40
42
44
46
48
Accuracy among all requests (%, 95% CI)
Pooled (45.2%)

**Supplementary Fig. 4 | Results by source dataset or program, and robustness to leaving programs or developers out.** A source dataset or program is the public dataset, registry or package from which a group of records came (NHANES, CDC PLACES, CDC WONDER, NCHS, ClinicalTrials.gov, or one of the three R meta-analysis packages). **a,** Best-estimate outcomes for each program, grouped as the three diagnostic R packages, the four population programs and ClinicalTrials.gov, with each program's estimation-family composition in its label. The programs differ in task mix (each contributes different estimation families), so this panel is not a quality ranking of sources. **b,** Accuracy after excluding one program at a time, with the questions each omission removes in its label; the vertical line is accuracy with all programs. This check asks whether one program alone produced the headline result. It does not imply that the remaining programs represent all clinical evidence. **c,** Developer robustness from the same reanalysis as Supplementary Table S50: accuracy among all quantity requests pooled, with equal weight per developer, and after leaving each developer out, with 95% parent-source-cluster bootstrap intervals.

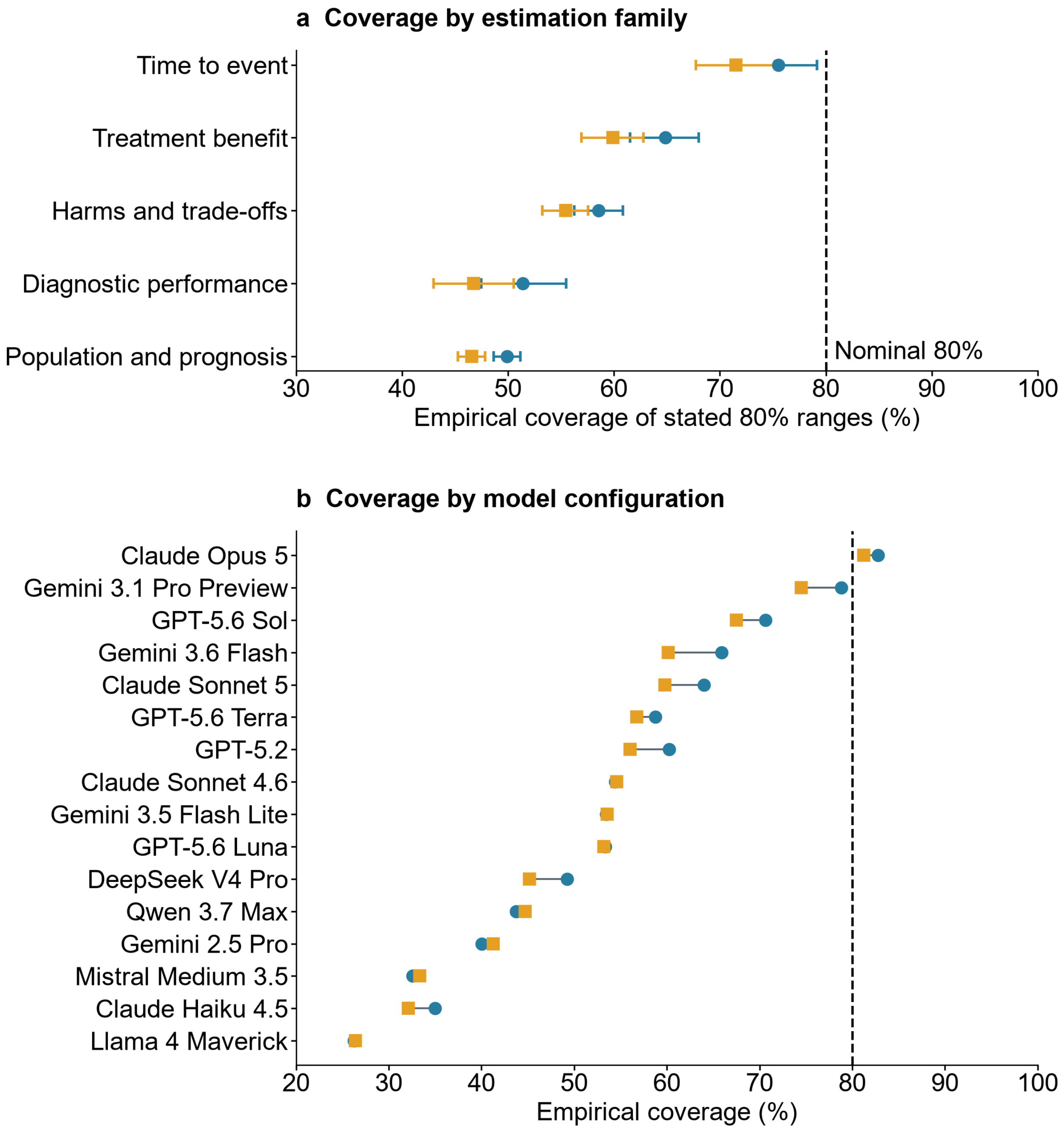


**Supplementary Fig. 5 | Calibration of stated 80% ranges by estimation family and configuration.** Panel a shows empirical coverage of stated 80% ranges per estimation family under both instruction policies, with 95% parent-source-cluster bootstrap intervals (10,000 replicates; the Wilson intervals remain in the source data), against the nominal 80% line. Panel b shows per-configuration point coverage under both policies, sorted by best-estimate coverage and drawn without intervals (the committed coverage table carries cluster bootstrap intervals at the family and overall grains only). Well-calibrated 80% ranges would cover approximately 80%; nearly all families and configurations fall short, and the deficit varies more by estimation family than by policy.

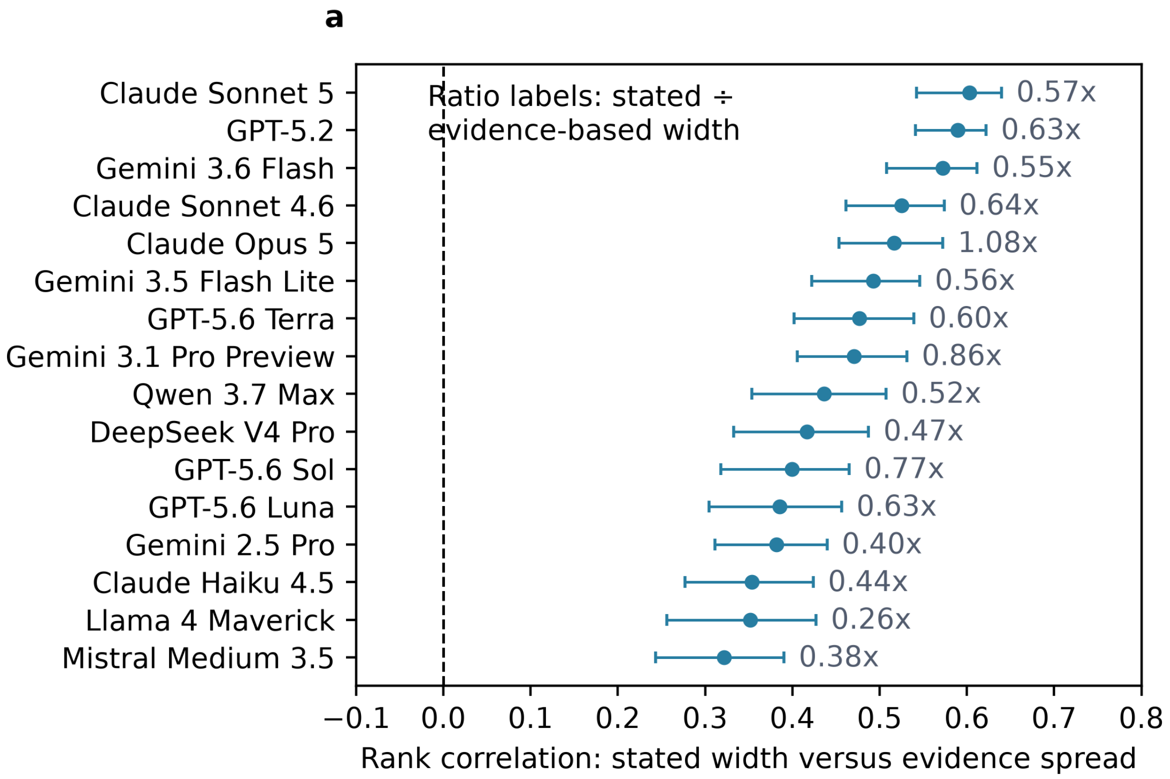
a
Ratio labels: stated ÷ evidence-based width
Claude Sonnet 5
GPT-5.2
Gemini 3.6 Flash
Claude Sonnet 4.6
Claude Opus 5
Gemini 3.5 Flash Lite
GPT-5.6 Terra
Gemini 3.1 Pro Preview
Qwen 3.7 Max
DeepSeek V4 Pro
GPT-5.6 Sol
GPT-5.6 Luna
Gemini 2.5 Pro
Claude Haiku 4.5
Llama 4 Maverick
Mistral Medium 3.5
0.57x
0.63x
0.55x
0.64x
1.08x
0.56x
0.60x
0.86x
0.52x
0.47x
0.77x
0.63x
0.40x
0.44x
0.26x
0.38x
−0.1
0.0
0.1
0.2
0.3
0.4
0.5
0.6
0.7
0.8
Rank correlation: stated width versus evidence spread

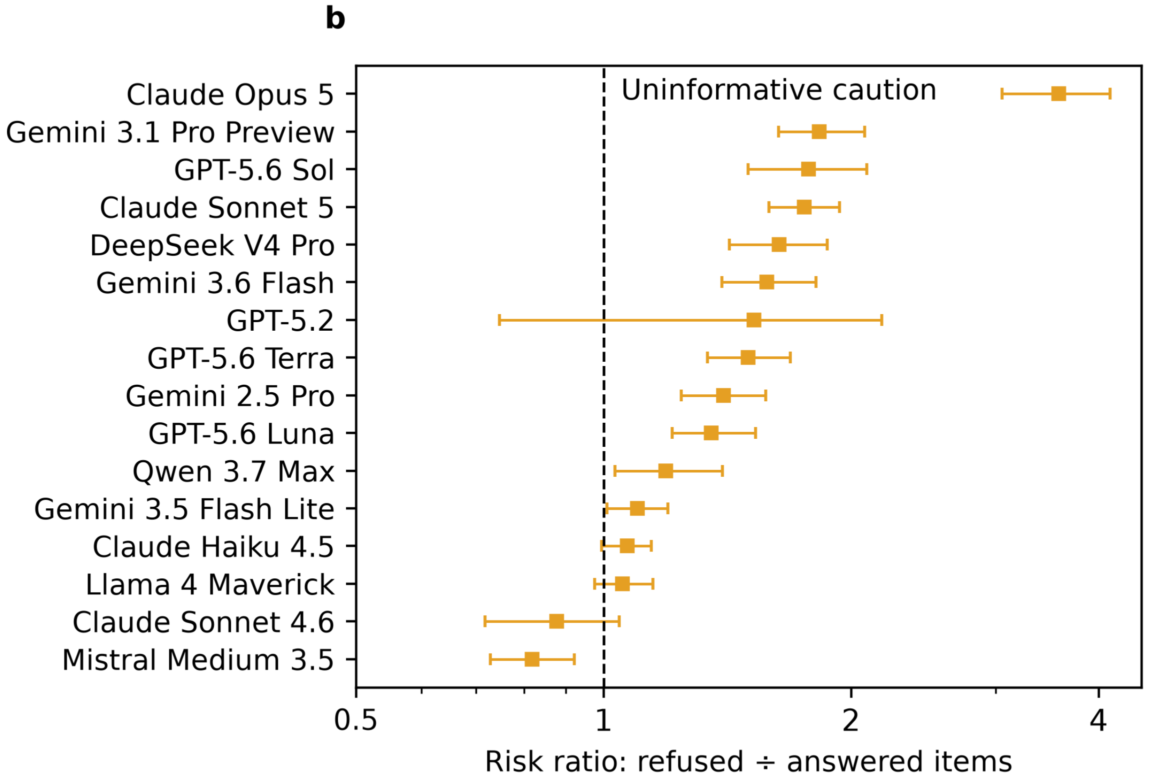
b
Uninformative caution
Claude Opus 5
Gemini 3.1 Pro Preview
GPT-5.6 Sol
Claude Sonnet 5
DeepSeek V4 Pro
Gemini 3.6 Flash
GPT-5.2
GPT-5.6 Terra
Gemini 2.5 Pro
GPT-5.6 Luna
Qwen 3.7 Max
Gemini 3.5 Flash Lite
Claude Haiku 4.5
Llama 4 Maverick
Claude Sonnet 4.6
Mistral Medium 3.5
0.5
1
2
4
Risk ratio: refused ÷ answered items

**Supplementary Fig. 6 | Stated widths track the evidence; caution quality varies fourfold.** Panel a shows, per model configuration, the quantity-stratified rank correlation between stated 80% range widths and the evidence-based predictive spread of the diagnostic questions' targets (the width of the target's central 80% interval; Supplementary Methods section 7) with source-cluster bootstrap 95% intervals; the annotations give the median ratio of stated width to evidence-based width. Panel b shows each configuration's caution risk ratio: the very-large-error rate of the quantities the configuration refused under answer-or-decline, evaluated from the same configuration's best-estimate answers to those quantities, divided by the very-large-error rate of the quantities it answered under answer-or-decline. A ratio above 1 means that refusals fell on the riskier quantities (informative caution), the dashed line at 1 marks uninformative caution, and a ratio below 1 means that refusals fell on the safer quantities (inverted caution). Human-readable model names are used; exact configuration identifiers are in Supplementary Table S4. Both analyses were added after the plan.

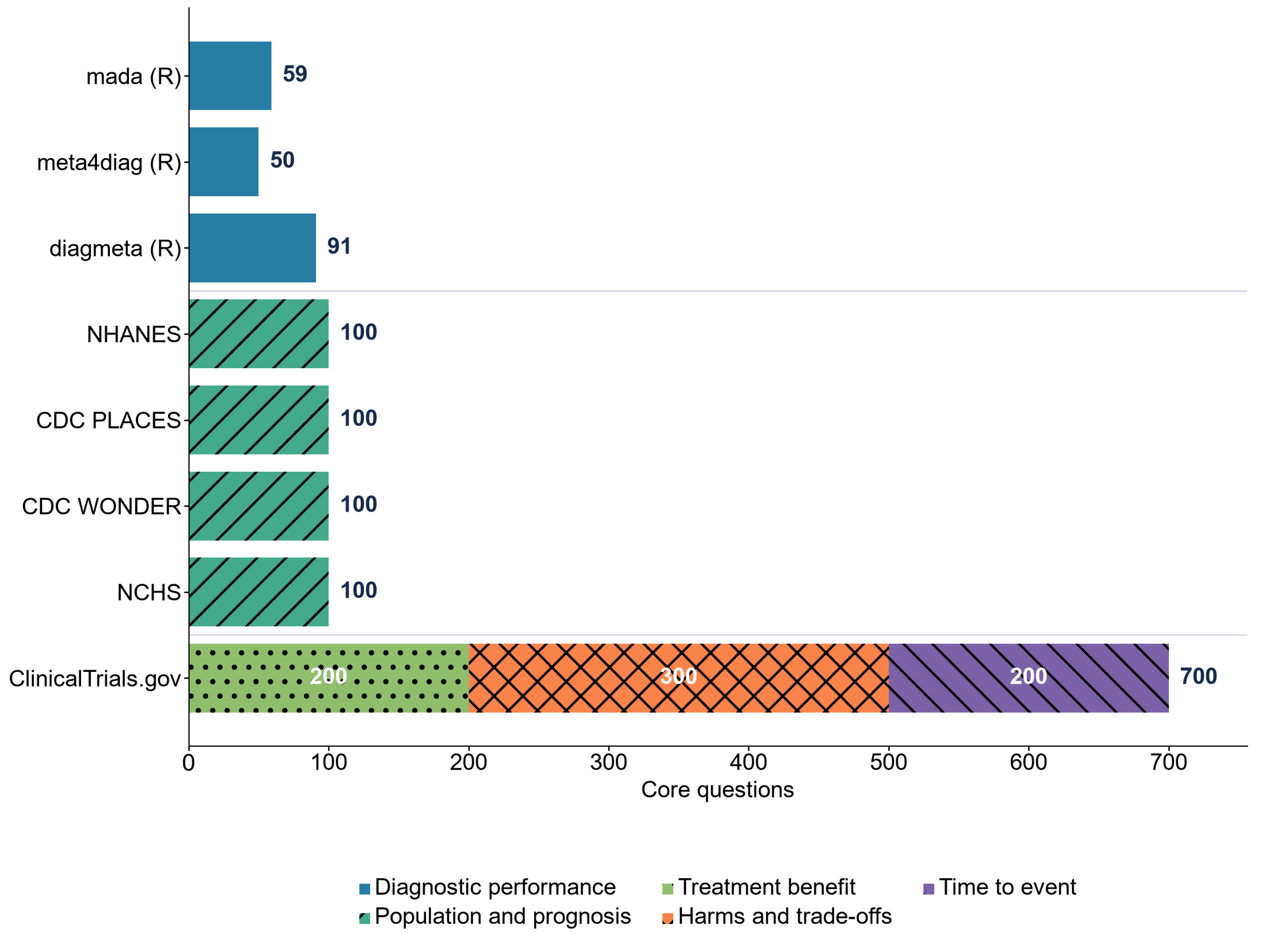


**Supplementary Fig. 7 | Composition of the final question set by source program and estimation family.** Counts of core questions are shown by data source and estimation family, with exact counts at segment and bar ends; programs are grouped as the three diagnostic R packages, the four population programs and ClinicalTrials.gov. The workbook provides the number of distinct sources and reference values. One source could contribute several questions or requested numbers, so these totals are not interchangeable. Confidence intervals kept observations from the same source together for this reason. The benchmark pipeline itself is Supplementary Figure 14.

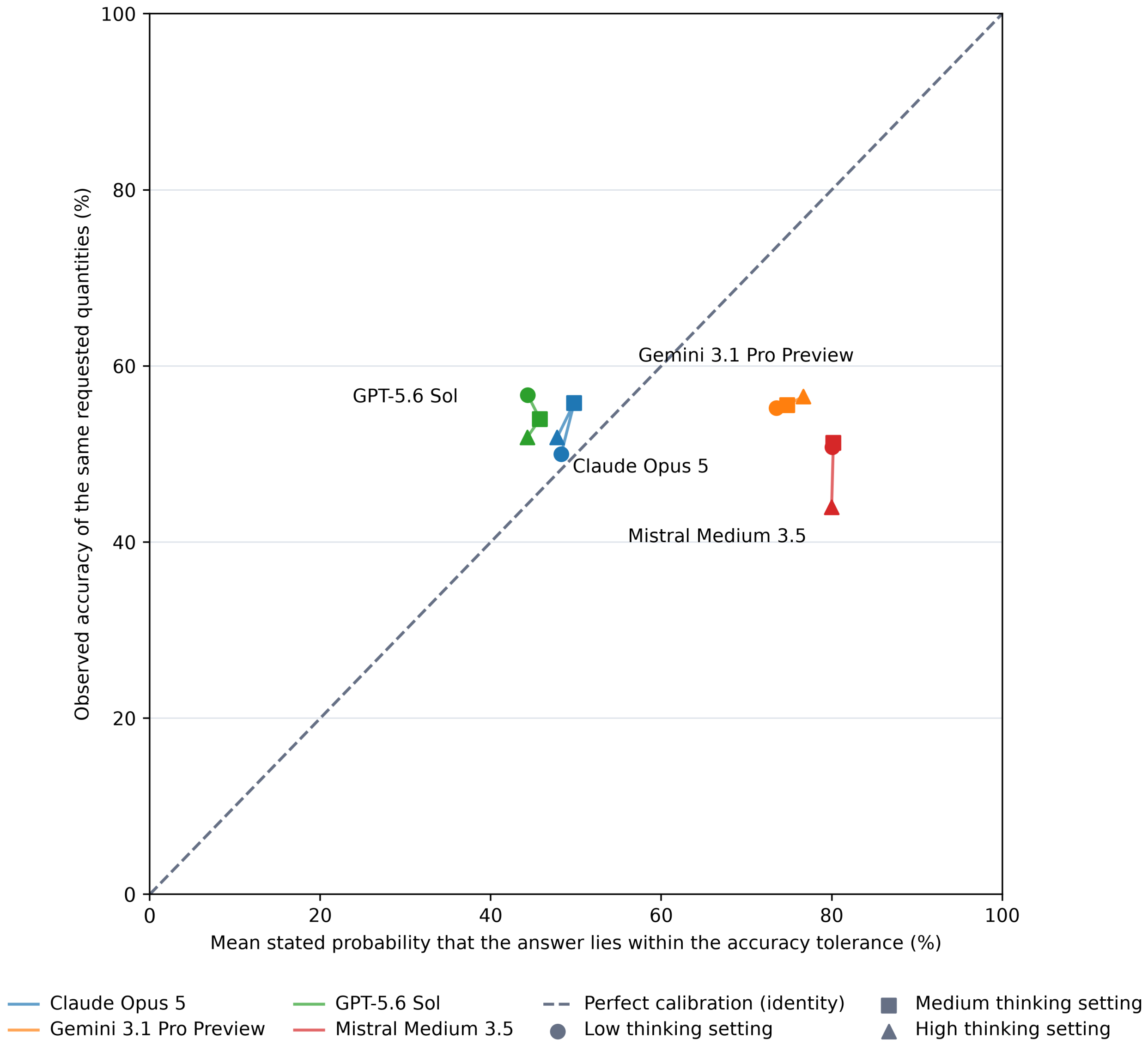


**Supplementary Fig. 8 | Numeric confidence was model-specific and stable across thinking doses.** For the four thinking-dose configurations at the low, medium and high provider-native reasoning settings, the mean stated probability that an answer lies within the accuracy tolerance (the schema field `probability_within_tolerance`, defined only by its name in the enforced schema and not described to the model; Supplementary Methods section 7, numeric confidence) is plotted against the observed accuracy of the same requested quantities; each point is one configuration at one setting, the marker shape gives the low, medium or high setting (key below the panel), the dashed identity line marks perfect calibration, and both axes span the full 0-100 range. The stated probabilities were computed among requested quantities for which the configuration stated a probability; Brier scores are in the source data.

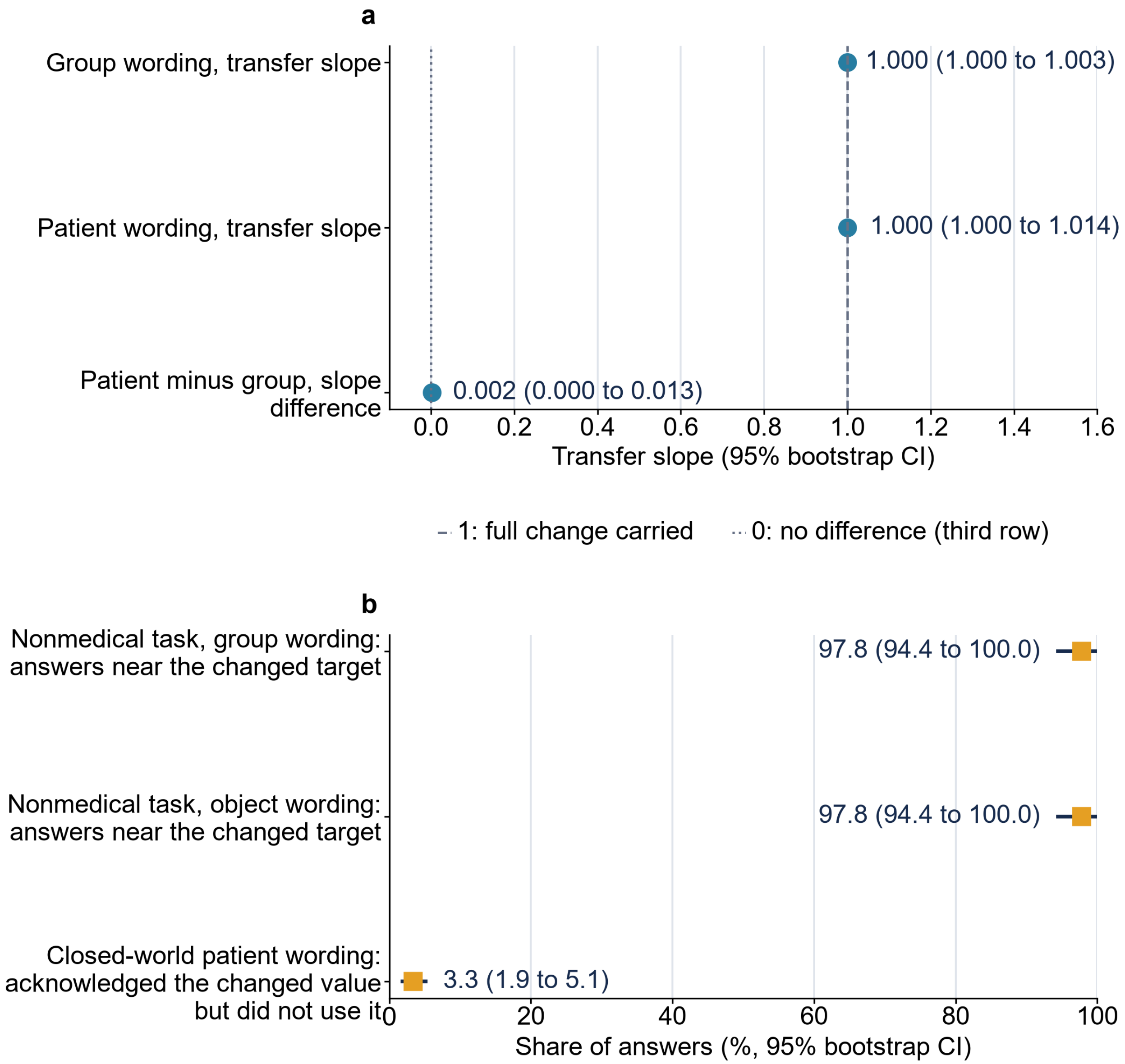
a
Group wording, transfer slope
1.000 (1.000 to 1.003)
Patient wording, transfer slope
1.000 (1.000 to 1.014)
Patient minus group, slope difference
0.002 (0.000 to 0.013)
0.0
0.2
0.4
0.6
0.8
1.0
1.2
1.4
1.6
Transfer slope (95% bootstrap CI)
1: full change carried
0: no difference (third row)
b
Nonmedical task, group wording: answers near the changed target
97.8 (94.4 to 100.0)
Nonmedical task, object wording: answers near the changed target
97.8 (94.4 to 100.0)
Closed-world patient wording: acknowledged the changed value but did not use it
3.3 (1.9 to 5.1)
0
20
40
60
80
100
Share of answers (%, 95% bootstrap CI)

**Supplementary Fig. 9 | Closed-world and nonmedical transfer in the patient-level experiment.** Prespecified secondary results with 95% source-cluster bootstrap intervals, from the closed-world check (30 invented disease–test families, every input supplied, five starting probabilities each) and the nonmedical control (the same 30 transformations as sensor-component problems). **a,** Closed-world transfer slopes: the slope of the change in the answer against the change in the correct value across the five starting probabilities, estimated through each family's middle target, under group wording and under patient wording, where 1 means the full change was carried; the third row is the patient-minus-group slope difference, whose reference is 0. **b,** Target-binding shares on a percentage axis, a different estimand from the slopes of panel a: the nonmedical target-binding rates under group wording and under individual-object wording, the share of answers within 0.05 of the changed target and closer to it than to the baseline target; and the share of closed-world patient-wording answers that acknowledged the changed value but did not use it (the protocol's "unbound after correct acknowledgement"), among responses whose acknowledgement field repeated the supplied starting probability exactly, which separates failing to register the input from failing to use it (the registered input was almost always used). The nonmedical rows are controls for the clinical result: the same arithmetic with the same inputs, outside medicine, was carried almost completely under both wordings (97.8%), so the clinical shortfall is not a general inability to perform the transformation. These control tasks supply the sensitivity, specificity and starting probability directly, so the matched clinical comparison is the repair arm that also supplies the inputs: it reached 78.8% of its 2,340 scheduled prompt pairs (Figure 3d), and the remaining gap to the nonmedical 97.8% is not closed by supplying the inputs alone. The identical printed values are coincidences of rounding and discreteness, not duplicated rows: the two transfer slopes differ before rounding (0.99993 and 1.00006, each with a lower bound just below 1), and the two nonmedical rates are separate analyses over 1,129 and 1,133 answers whose configuration-level shares coincide exactly.

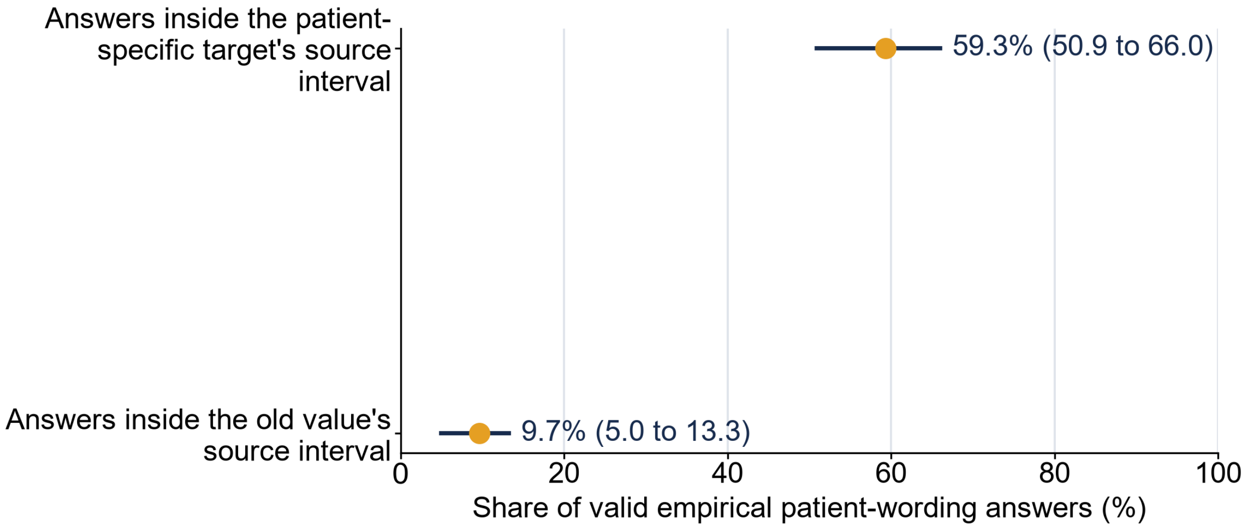


**Supplementary Fig. 10 | Source-uncertainty check for the empirical module.** Among valid empirical patient-wording answers at the changed value, the share that fell inside the survey confidence interval of the patient-specific value (59.3%) and inside the interval of the old value (9.7%), with 95% source-cluster bootstrap intervals; the axis names this denominator. The two compatibilities are distinct: the first asks whether an answer is consistent with the patient-specific estimate given its survey uncertainty, the second whether it is still consistent with the old (population) estimate.

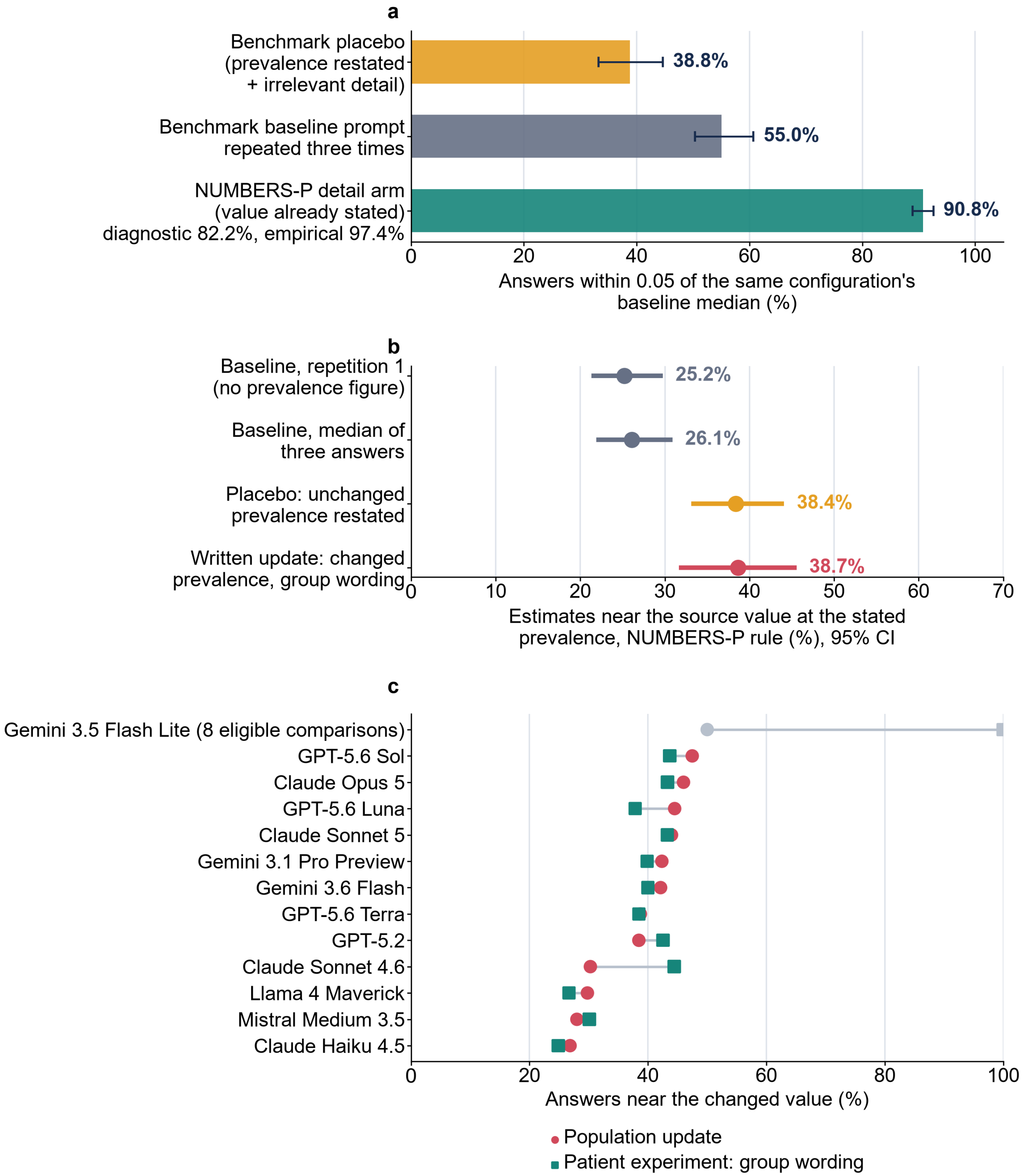

a
Benchmark placebo (prevalence restated + irrelevant detail)
Benchmark baseline prompt repeated three times
NUMBERS-P detail arm (value already stated) diagnostic 82.2%, empirical 97.4%
38.8%
55.0%
90.8%
0
20
40
60
80
100
Answers within 0.05 of the same configuration's baseline median (%)
b
Baseline, repetition 1 (no prevalence figure)
Baseline, median of three answers
Placebo: unchanged prevalence restated
Written update: changed prevalence, group wording
25.2%
26.1%
38.4%
38.7%
0
10
20
30
40
50
60
70
Estimates near the source value at the stated prevalence, NUMBERS-P rule (%), 95% CI
c
Gemini 3.5 Flash Lite (8 eligible comparisons)
GPT-5.6 Sol
Claude Opus 5
GPT-5.6 Luna
Claude Sonnet 5
Gemini 3.1 Pro Preview
Gemini 3.6 Flash
GPT-5.6 Terra
GPT-5.2
Claude Sonnet 4.6
Llama 4 Maverick
Mistral Medium 3.5
Claude Haiku 4.5
0
20
40
60
80
100
Answers near the changed value (%)
Population update
Patient experiment: group wording

**Supplementary Fig. 11 | Cross-study reconciliation of prevalence-restatement controls and group-wording updates (exploratory reanalysis of existing responses; no new model calls). a,** Stability, defined throughout as an answer within 0.05 of the median of the same configuration's three baseline answers for the item, with the cells of the prevalence-update experiment (2,300 eligible cells) or the items of the patient-level experiment as denominators: the benchmark placebo answer (unchanged prevalence restated as a number plus an irrelevant scheduling detail) against the median of the three base answers, 38.8%; the three benchmark base answers themselves all within 0.05 of one another, 55.0%; and the patient-level irrelevant-detail arm against the median of the three patient-wording baseline answers, 90.8% of the 6,637 valid answers with a baseline median (95% CI, 88.9–92.6; 82.2% diagnostic, 97.4% empirical). The second bar is a different contrast from the first: it measures the dispersion of the base answers with no sentence added, and is shown as the reference level against which the placebo's 38.8% is read, since an added sentence cannot be expected to leave more answers in place than the base prompt's own repeats do. **b,** Benchmark updating arms scored by the near-target rule of the patient-level experiment against source values: an answer counts when it is within 0.05 of the source value at the stated prevalence and closer to it than to the value at the other prevalence, among valid answers. **c,** Group-wording update at the changed value across the 13 shared configurations, scored by the same source rule: the benchmark written arm and the patient-level group-wording arm at the changed value (diagnostic items); the configuration with 8 eligible comparisons is grey. Error bars show 95% source-cluster bootstrap intervals (10,000 replicates). No prespecified outcome changed.

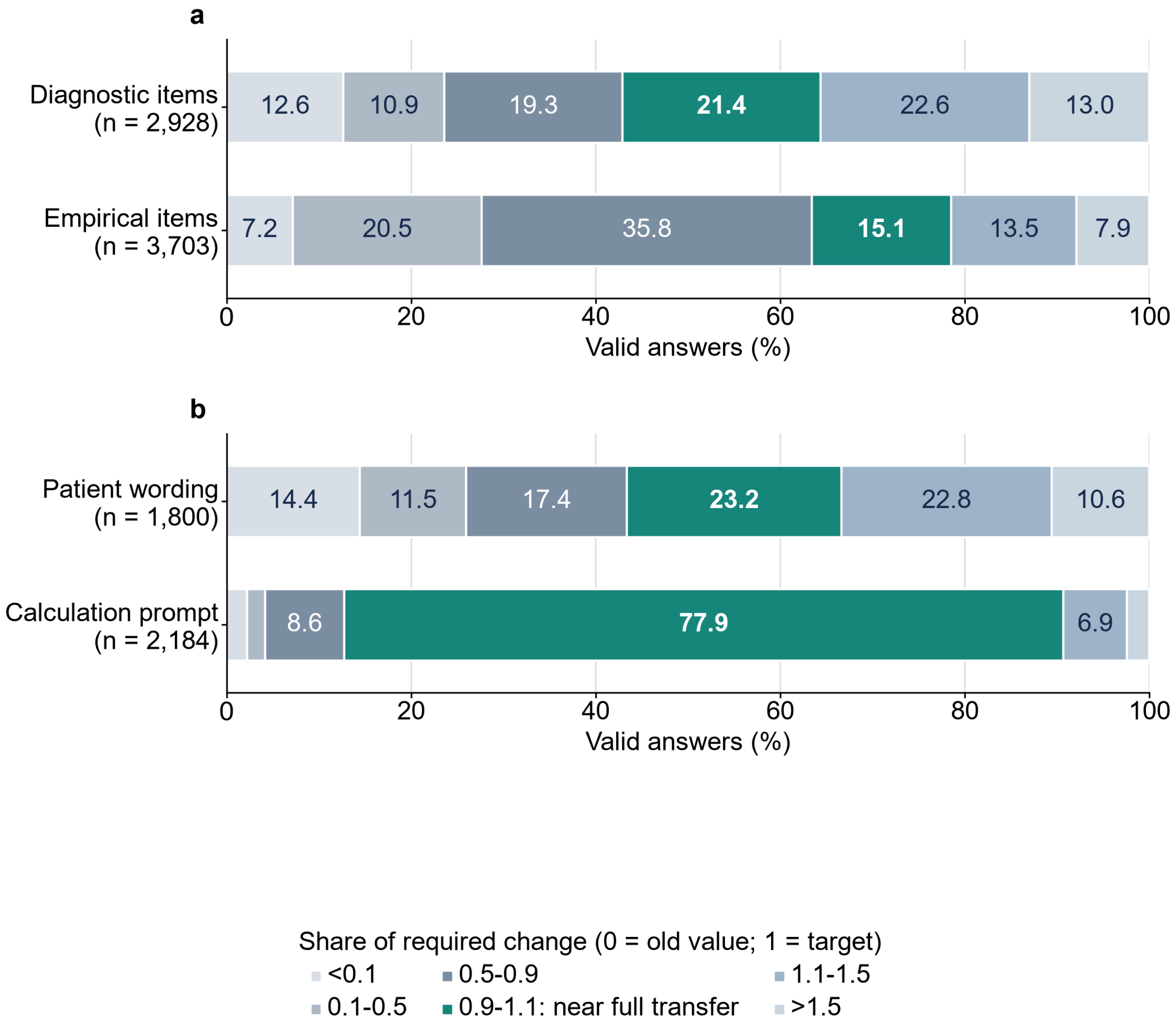
a
Diagnostic items (n = 2,928)
12.6
10.9
19.3
21.4
22.6
13.0
Empirical items (n = 3,703)
7.2
20.5
35.8
15.1
13.5
7.9
0
20
40
60
80
100
Valid answers (%)
b
Patient wording (n = 1,800)
14.4
11.5
17.4
23.2
22.8
10.6
Calculation prompt (n = 2,184)
8.6
77.9
6.9
0
20
40
60
80
100
Valid answers (%)
Share of required change (0 = old value; 1 = target)
<0.1
0.5-0.9
1.1-1.5
0.1-0.5
0.9-1.1: near full transfer
>1.5

**Supplementary Fig. 12 | Where patient-level answers landed between the old and the patient-specific value.** Each valid answer is placed by the share of the required change it carried, (answer − old value)/(patient-specific value − old value): 0 is the old (population) value and 1 the patient-specific value. **a,** Patient-wording answers at the changed value for the diagnostic and empirical modules, restricted to substitution-eligible items (those whose two values differ by at least 0.075, as in the prespecified primary analysis); the medians of 0.99 and 0.75 are medians over all pooled valid answers, whereas the Article's 81.4% is the median of equally weighted configuration-by-module summaries, so they are different summaries of the same movement variable. **b,** The 60 diagnostic repair items: the same answers under patient wording and the calculated answers after the prompt that supplied the inputs and asked for the calculation. The highlighted 0.9–1.1 bin is near full transfer, a share of the required change between 0.9 and 1.1 (23.2% before, 77.9% after; median 1.00 after); the bin is an interval, not literal equality with the target (exact target, which this analysis does not measure). Each bar stacks the six bins to 100% of its valid answers, so the bins read as ordered categories: segment length encodes only the share of answers, not the bins' unequal numeric widths, and segments below 5% carry no printed value on the plate (Supplementary Table S43b holds every count). Near full transfer is a different classification from the prespecified near-target rule used for the main repair result (within 0.05 of the patient-specific value, the Article's case-conditioned target, and closer to it than to the old value; 29.7% to 78.8%). Percentages are of valid answers; n per bar in its label (Supplementary Tables S43a and S43b).

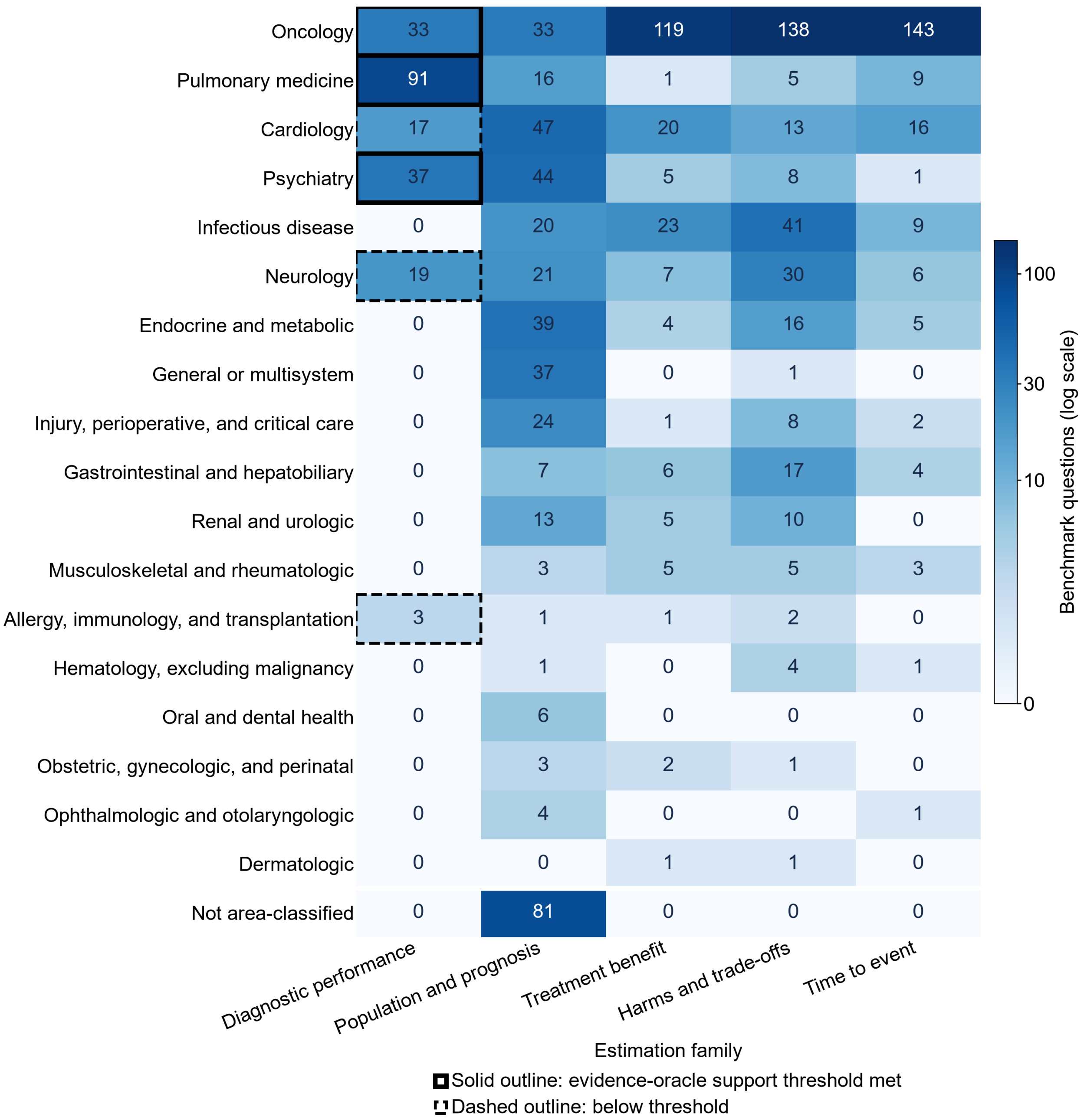

Oncology
Pulmonary medicine
Cardiology
Psychiatry
Infectious disease
Neurology
Endocrine and metabolic
General or multisystem
Injury, perioperative, and critical care
Gastrointestinal and hepatobiliary
Renal and urologic
Musculoskeletal and rheumatologic
Allergy, immunology, and transplantation
Hematology, excluding malignancy
Oral and dental health
Obstetric, gynecologic, and perinatal
Ophthalmologic and otolaryngologic
Dermatologic
Not area-classified
33 33 119 138 143
91 16 1 5 9
17 47 20 13 16
37 44 5 8 1
0 20 23 41 9
19 21 7 30 6
0 39 4 16 5
0 37 0 1 0
0 24 1 8 2
0 7 6 17 4
0 13 5 10 0
0 3 5 5 3
3 1 1 2 0
0 1 0 4 1
0 6 0 0 0
0 3 2 1 0
0 4 0 0 1
0 0 1 1 0
0 81 0 0 0
Diagnostic performance
Population and prognosis
Treatment benefit
Harms and trade-offs
Time to event
Estimation family
Benchmark questions (log scale)
100
30
10
0
Solid outline: evidence-oracle support threshold met
Dashed outline: below threshold

**Supplementary Fig. 13 | Coverage of the 18 clinical areas by estimation family.** Each cell gives the number of benchmark questions for one clinical area in one estimation family, on a logarithmic colour scale with the count printed. Row labels use the Article's specialty names where the text names one (pulmonary medicine, psychiatry, oncology, neurology, cardiology); the other rows keep the frozen clinical-area taxonomy names, the data files keep the taxonomy names throughout, and the key beneath the grid decodes the solid and dashed outlines. The evidence-oracle comparison of Figure 1a exists only for the diagnostic family, because the oracle refits a diagnostic meta-analysis without the withheld study; the three outlined diagnostic cells are the specialties whose eight diagnostic quantities meet the support threshold of at least 30 questions and 10 parent-source clusters per quantity cell (pulmonary medicine, psychiatry, oncology), the dashed cells sit below it (neurology and cardiology, which appear in Figure 1a as hollow markers, and allergy with three questions), and the remaining specialties hold no diagnostic meta-analyses. The other families reach most specialties: harm questions sit in 16 of the 18, population questions in 17, treatment-benefit questions in 14 and time-to-event questions in 12 (Supplementary Table S46 lists the diagnostic support per specialty). The bottom grid row counts the 81 population questions whose sources carry no clinical-area label; with that row, every column sums to its family total and the grid covers all 1,300 benchmark questions.

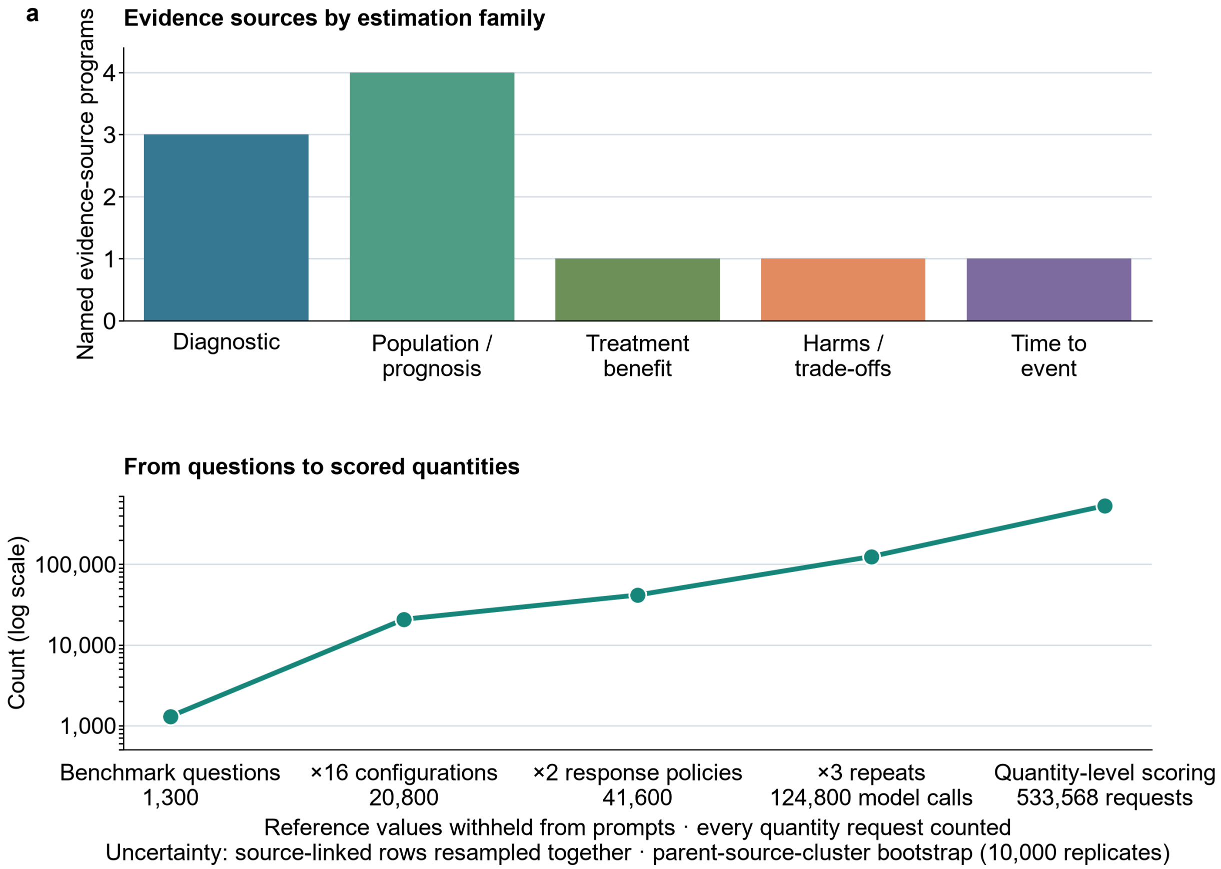


**Supplementary Figure 14 | Benchmark construction and evaluation design.** **a,** Evidence-source programs supporting the five estimation families, named as in the Methods. **b,** The factorial design: 1,300 clinical questions × 16 model configurations × 2 response policies × 3 repeats produced 124,800 model calls. Quantity-level scoring yielded 533,568 requests because a question could ask for more than one number. Reference values were withheld from the prompts, every requested quantity was counted, and source-linked rows were kept together in the parent-source-cluster bootstrap (10,000 replicates). The four studies were designed sequentially, and each was frozen before its own model calls (Supplementary Table S42); later reanalyses used existing responses and made no new model call. The composition of the question set by source program is Supplementary Fig. 7.

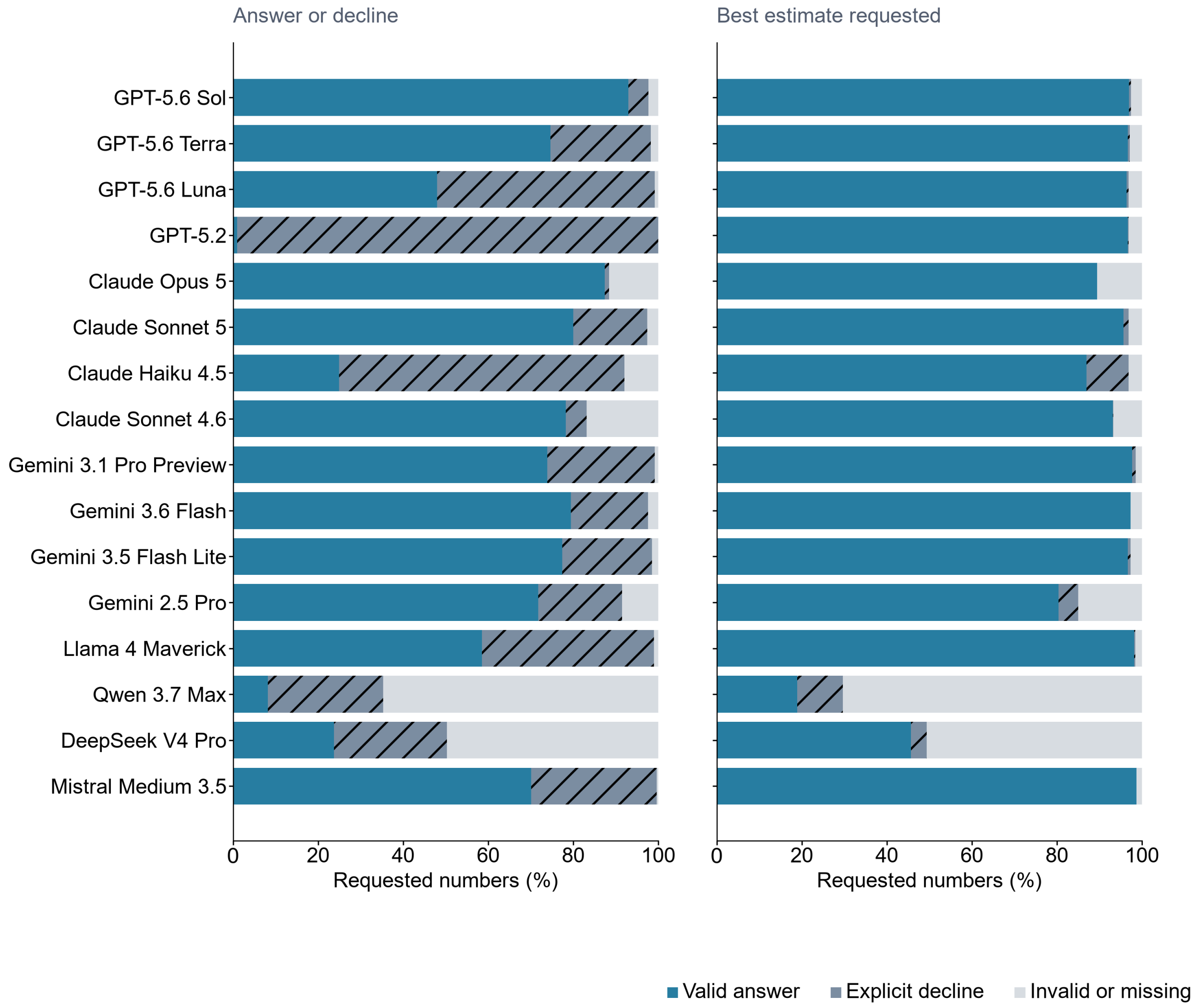


**Supplementary Figure 15 | What happened to every requested number.** Each bar separates valid answers, explicit declines, and invalid or missing responses for one model and instruction, in the fixed configuration order of Supplementary Figure 24. Every bar includes all requested numbers. Hatched segments indicate explicit declines: the model chose not to provide an estimate and gave a permitted reason. The invalid-or-missing group combines several technical failure types that are listed separately in Supplementary Tables S7a and S7b. Model-level median valid-answer rates were 72.8% (IQR, 42.2–78.5) under answer-or-decline and 96.5% (88.9–97.0) under best-estimate. GPT-5.2's large nonanswer share under answer-or-decline consists mostly of explicit declines rather than malformed JSON.

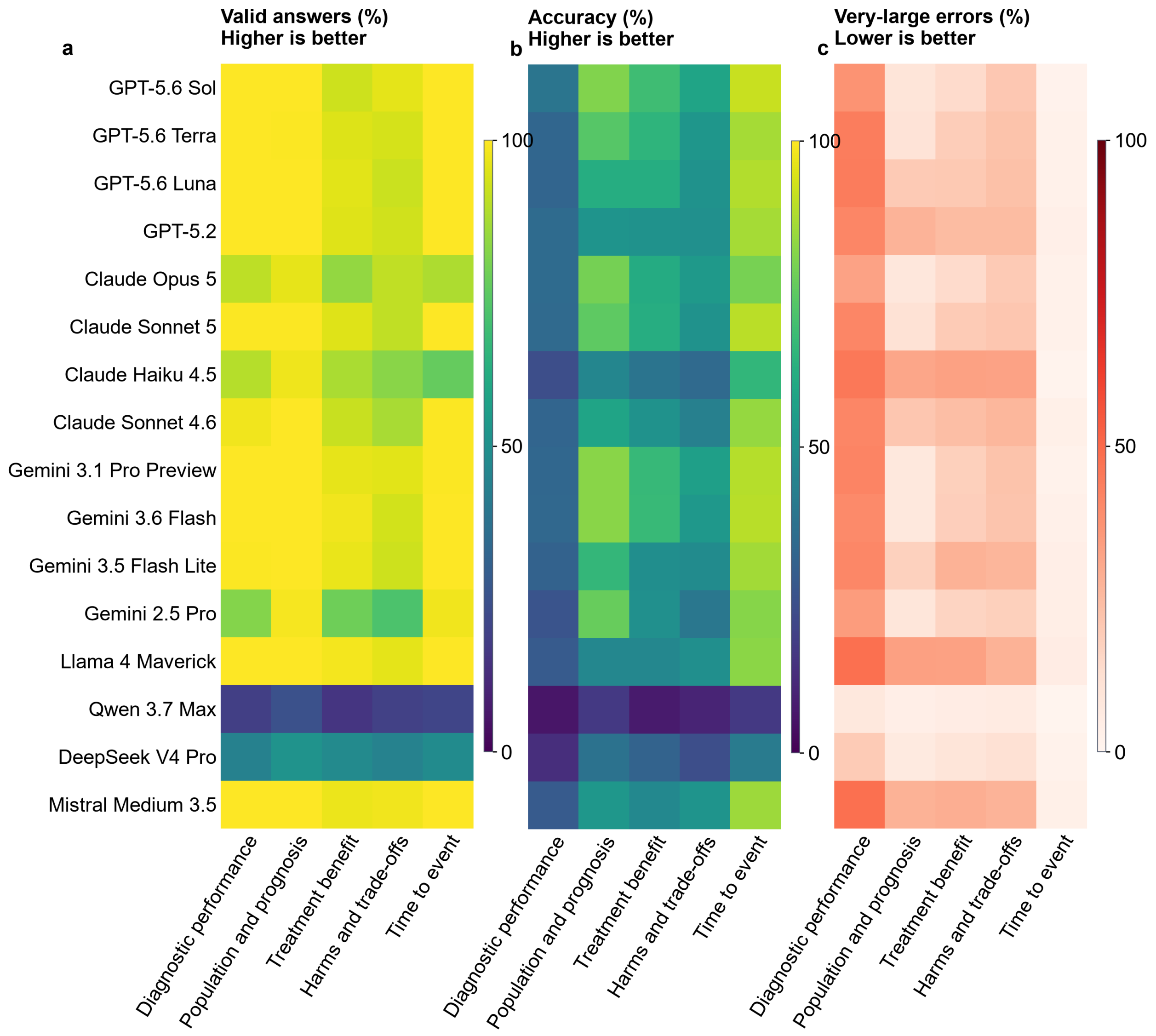


**Supplementary Figure 16 | Best-estimate results for every model and estimation family. a,** Percentage of requests receiving a valid number. **b,** Percentage accurate among all requests. **c,** Percentage with very-large errors. The valid-answer and accuracy grids use a blue-green scale on which higher is better; the very-large-error grid uses a red scale on which lower is better, so a bright square is not favourable in every grid. Model order was fixed before results and is not a ranking. These three percentages should be read together. Answering often does not guarantee reliable estimates.

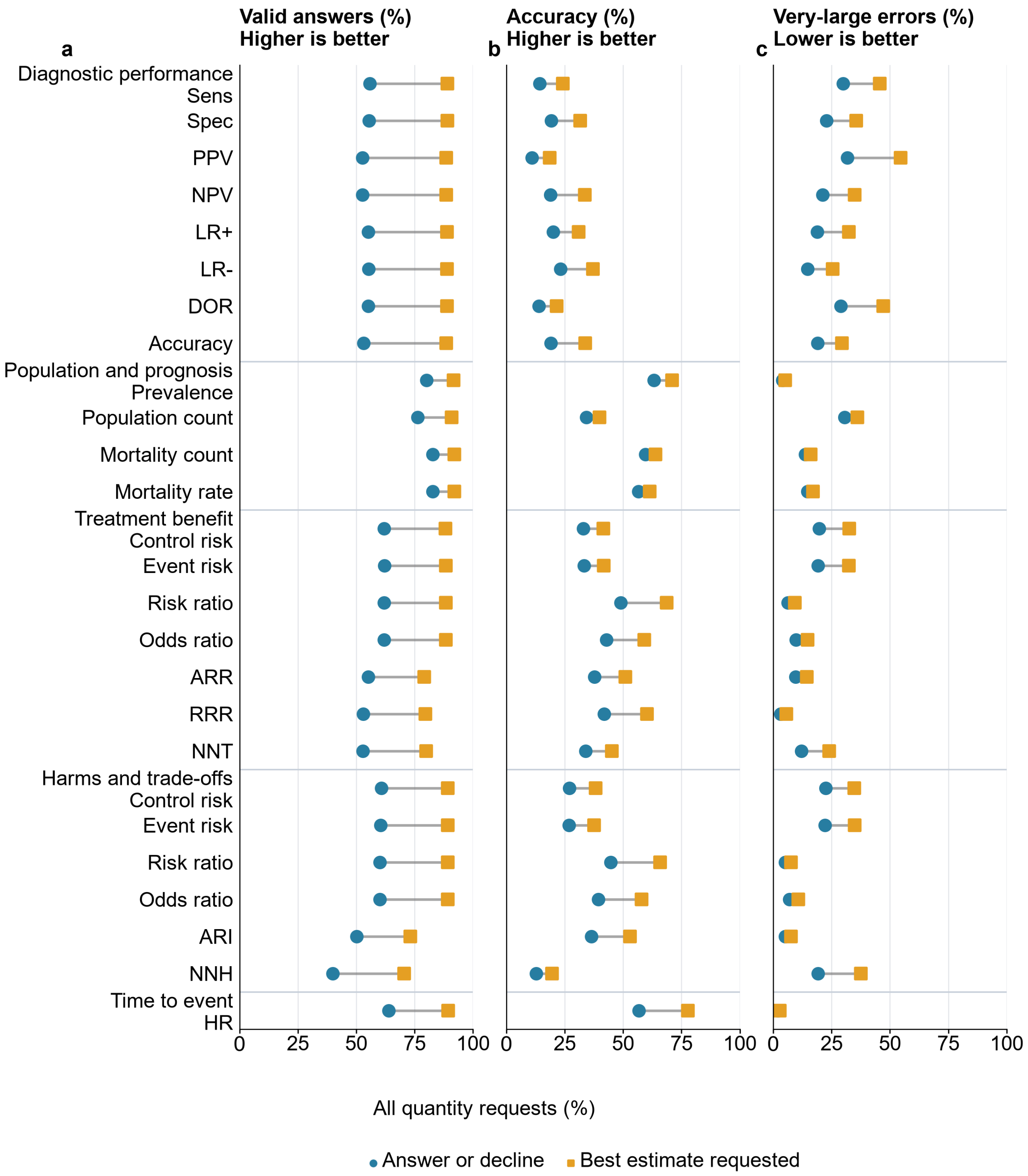

a
Valid answers (%)
Higher is better
b
Accuracy (%)
Higher is better
c
Very-large errors (%)
Lower is better
Diagnostic performance
Sens
Spec
PPV
NPV
LR+
LR-
DOR
Accuracy
Population and prognosis
Prevalence
Population count
Mortality count
Mortality rate
Treatment benefit
Control risk
Event risk
Risk ratio
Odds ratio
ARR
RRR
NNT
Harms and trade-offs
Control risk
Event risk
Risk ratio
Odds ratio
ARI
NNH
Time to event
HR
0 25 50 75 100
All quantity requests (%)
Answer or decline
Best estimate requested

**Supplementary Figure 17 | Results for all 26 clinical measures under both instructions. a,** Valid-answer percentage. **b,** Source-result accuracy. **c,** Very-large-error percentage. For each measure, one dot per instruction policy shows the result among all quantity requests, and the two dots are joined only within that measure; the measures are grouped under the estimation-family headings used in Supplementary Figure 24, in the fixed taxonomy order. Looking at each measure prevents averages for a broad estimation family from hiding difficult tasks such as predictive values, risk differences and numbers needed to treat or harm. It also shows whether a change between instructions occurred throughout an estimation family or in only a few measures. Sens, sensitivity; Spec, specificity; PPV and NPV, positive and negative predictive value; LR+ and LR−, positive and negative likelihood ratio; DOR, diagnostic odds ratio; ARR, absolute risk reduction; RRR, relative risk reduction; NNT, number needed to treat; ARI, absolute risk increase; NNH, number needed to harm; HR, hazard ratio.

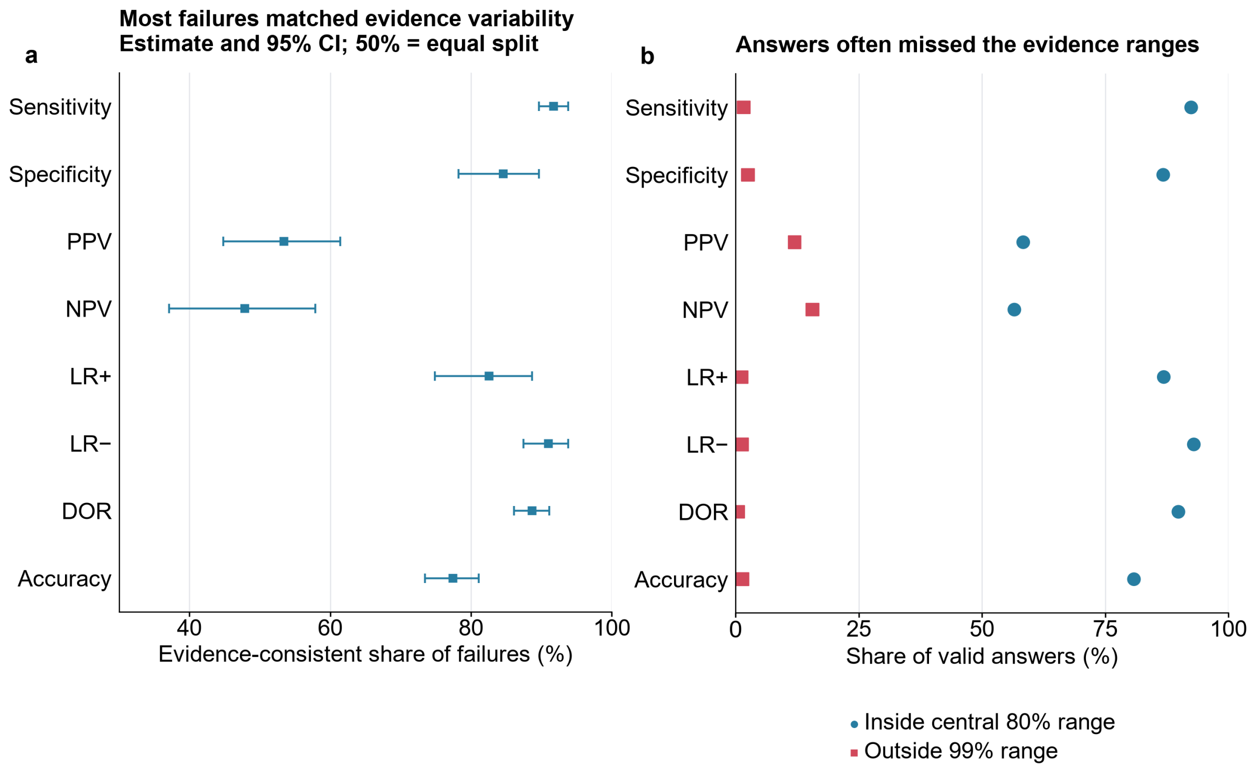


**Supplementary Figure 18 | Why diagnostic answers failed: evidence variability versus model error. a,** The share of prespecified tolerance failures consistent with the evidence's predictive range, by diagnostic measure, with cluster-bootstrap 95% CIs; the remaining share is model displacement, and 50% is an equal split. **b,** Among all valid answers, the share inside the evidence's central 80% range and the share outside its 99% range. The inside share rose with the number $k$ of other studies in the meta-analysis: 68.2% at $k \leq 9$, 74.6% at $k$ = 10–24 and 83.2% at $k \geq 25$ (77.6% restricted to $k \geq 10$). This secondary rescoring was added after the benchmark results were known; the prespecified tolerance results are unchanged.

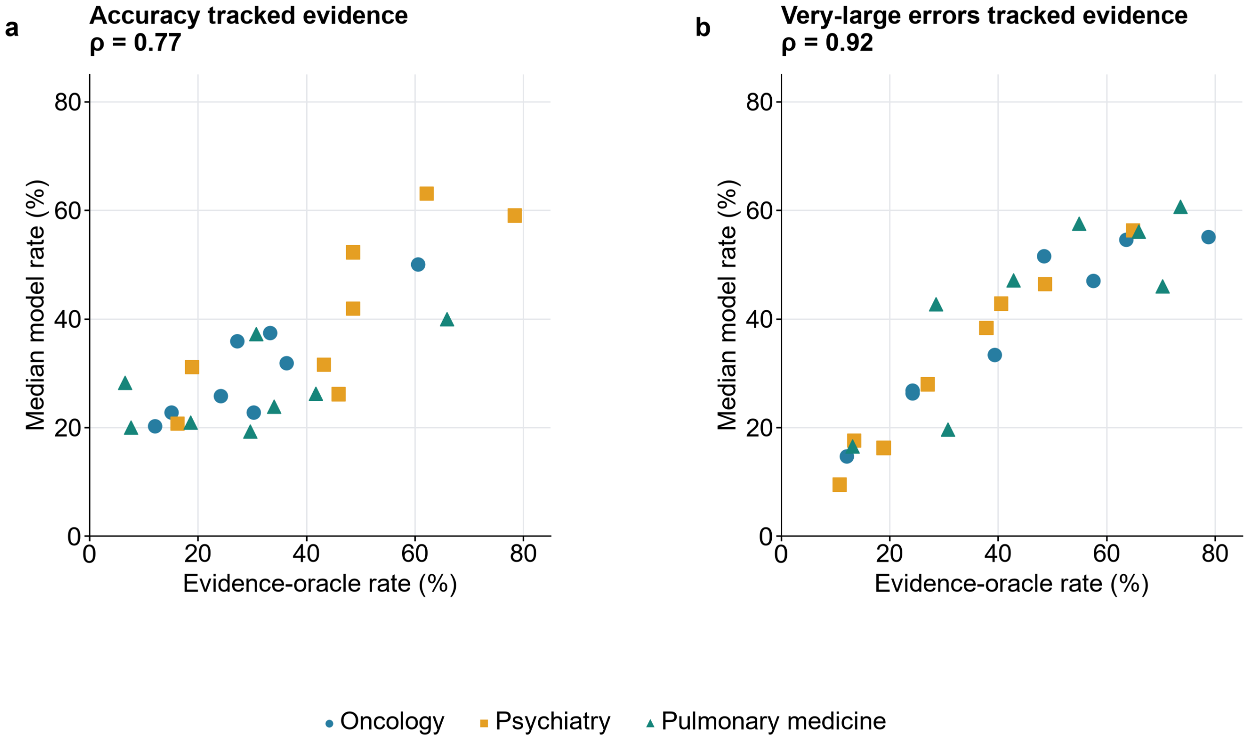


**Supplementary Figure 19 | Clinical-area differences in diagnostic performance tracked evidence predictability.** Each point is one clinical-area-by-quantity combination meeting the fixed support threshold of at least 30 questions and 10 parent-source clusters; colours identify oncology, psychiatry and pulmonary medicine. Model values are medians across 16 configurations under the best-estimate instruction. The x-axis is the accuracy (**a**) or very-large-error rate (**b**) of the response-independent within-meta-analysis evidence oracle. Across the 24 supported combinations, rank correlations were 0.77 for accuracy and 0.92 for very-large-error rate. This post hoc analysis describes benchmark composition and does not estimate general specialty competence.

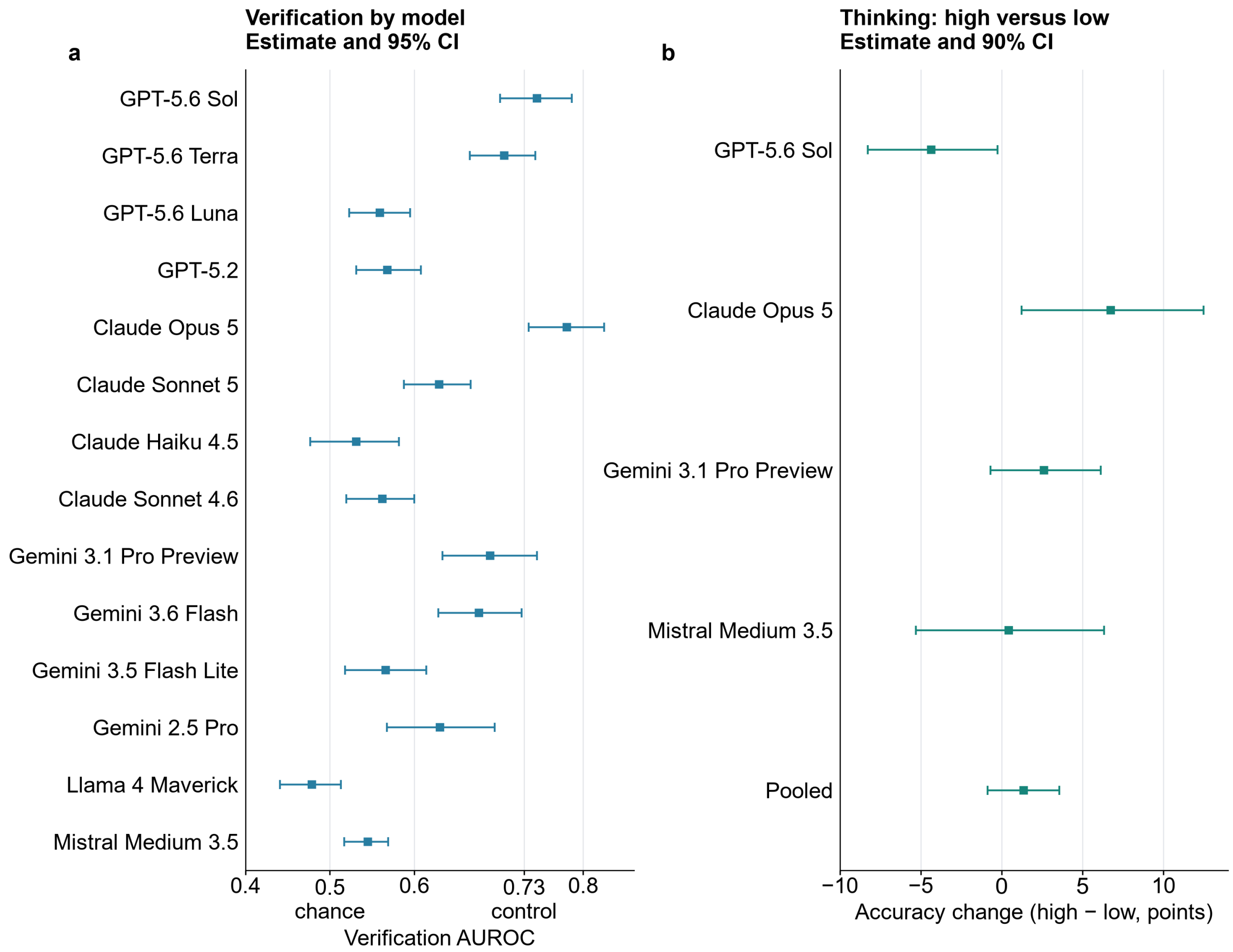


**Supplementary Figure 20 | Verification controls and the thinking-dose pilot. a,** Verification AUROC and 95% CI for 14 configurations in fixed order. The axis marks chance at 0.5 and the candidate-value-only control at 0.730, which was reached without any model, so model verification results remain descriptive; per-configuration AUROCs ran from 0.479 to 0.780. **b,** High-minus-low change in accuracy with two-sided 90% intervals. The pooled contrast of the 1,200-call pilot over 1,840 paired quantity requests was 1.4 percentage points (two-sided 90% interval, −0.9 to 3.6), and configuration changes ran from −4.3 to +6.7 points. Low, medium and high are provider-native settings, not a common reasoning-dose scale across providers.

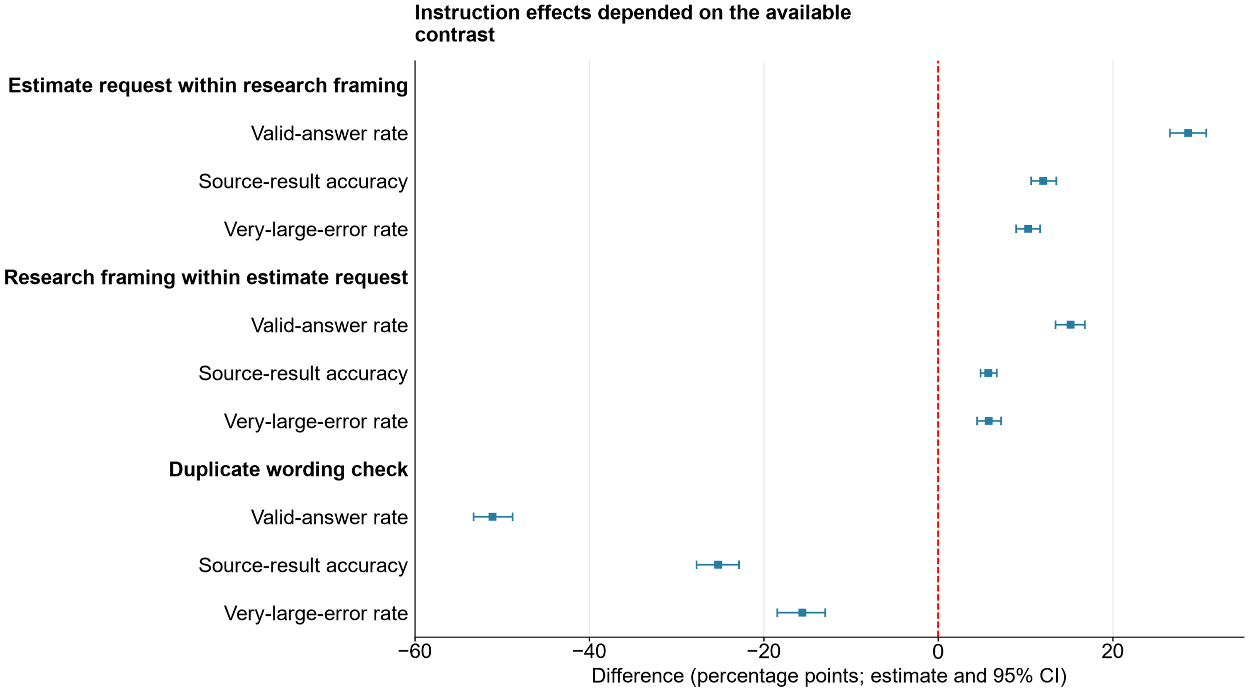


**Supplementary Figure 21 | Estimable instruction contrasts and the duplicate-wording check.** The planned 2 × 2 design (research framing × estimate request) could not identify the interaction because one factor combination was never run. That cell was never scheduled: the frozen amendment added only two system messages to the benchmark's two policies, and every benchmark policy message carries the research frame. Each named contrast shows the difference in valid answers, source-result accuracy and very-large errors in percentage points with 95% CIs; higher very-large-error differences are worse. An estimate request within the research frame raised valid answers, source-result accuracy and very-large errors by 28.6, 12.0 and 10.3 percentage points, and research framing within an estimate request raised them by 15.2, 5.8 and 5.8 points. The final group shows the large difference between nominally duplicate wordings.

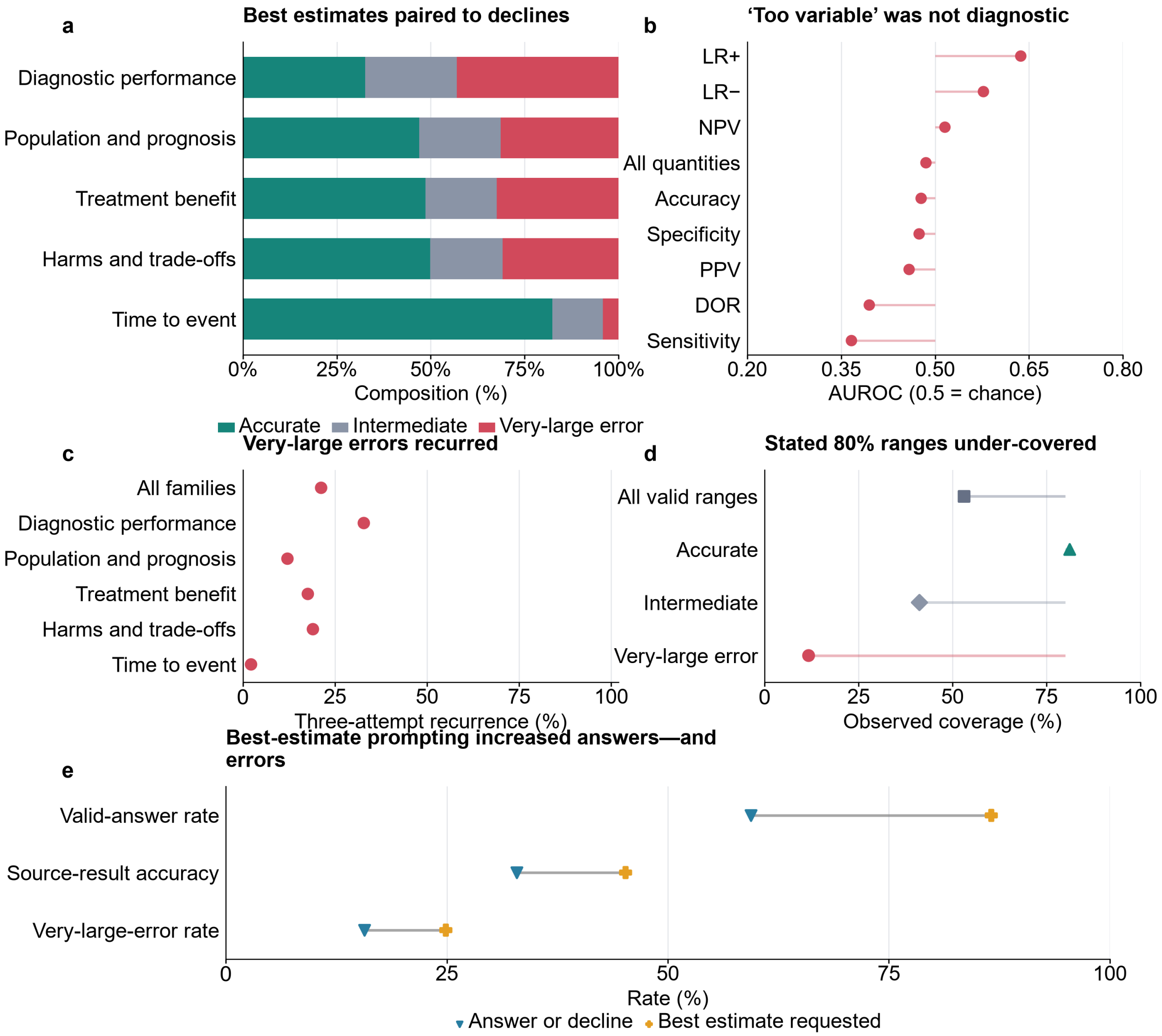
a
Best estimates paired to declines
Diagnostic performance
Population and prognosis
Treatment benefit
Harms and trade-offs
Time to event
0%
25%
50%
75%
100%
Composition (%)
Accurate
Intermediate
Very-large error
b
'Too variable' was not diagnostic
LR+
LR−
NPV
All quantities
Accuracy
Specificity
PPV
DOR
Sensitivity
0.20
0.35
0.50
0.65
0.80
AUROC (0.5 = chance)
c
Very-large errors recurred
All families
Diagnostic performance
Population and prognosis
Treatment benefit
Harms and trade-offs
Time to event
0
25
50
75
100
Three-attempt recurrence (%)
d
Stated 80% ranges under-covered
All valid ranges
Accurate
Intermediate
Very-large error
0
25
50
75
100
Observed coverage (%)
e
Best-estimate prompting increased answers—and errors
Valid-answer rate
Source-result accuracy
Very-large-error rate
0
25
50
75
100
Rate (%)
Answer or decline
Best estimate requested

**Supplementary Figure 22 | Visible caution, repetition and ordinary prompting did not reliably repair or expose the failure. a,** Outcomes among the 67,718 usable best estimates paired to declines, by estimation family: 44.1% were accurate and 34.9% were very-large errors overall; for diagnostic refusals, 32.6% and 43.0%. **b,** Whether "too variable to estimate" separated questions with more variable evidence: pooled AUROC, 0.485 (95% bootstrap CI, 0.453–0.520; $n$ = 261), where 0.5 is chance. Across configurations, refused-versus-answered very-large-error risk ratios ranged from 3.6 to 0.82. **c,** Very-large errors on all three attempts, pooled and by estimation family. They recurred in 21.2% of 69,746 groups with all three answers valid, from 88,928 scheduled groups, and 99.1% kept the same direction; configuration rates ranged from 12.1% to 31.4%. **d,** Coverage of stated 80% ranges: 53.1% across 220,006 valid ranges; 81.2%, 41.2% and 11.7% when the point estimate was accurate, intermediate or a very-large error. Under answer-or-decline and best-estimate, family coverage was 75.5% and 71.5% for time to event, 51.4% and 46.8% for diagnostic performance, and 49.9% and 46.6% for population and prognosis. **e,** The two response policies over 266,784 matched requests per policy. Best-estimate prompting changed valid-answer, source-result accuracy and very-large-error rates by +27.2, +12.3 and +9.2 percentage points (95% parent-source-cluster bootstrap CIs, 26.4–28.1, 11.7–12.9 and 8.7–9.8). Among 150,516 requests answered under both policies, accuracy was 55.4% and 55.9%, and very-large-error rates were 26.4% and 26.0%. Transitions: Table S11; policy messages: Supplementary Methods section 2.

a

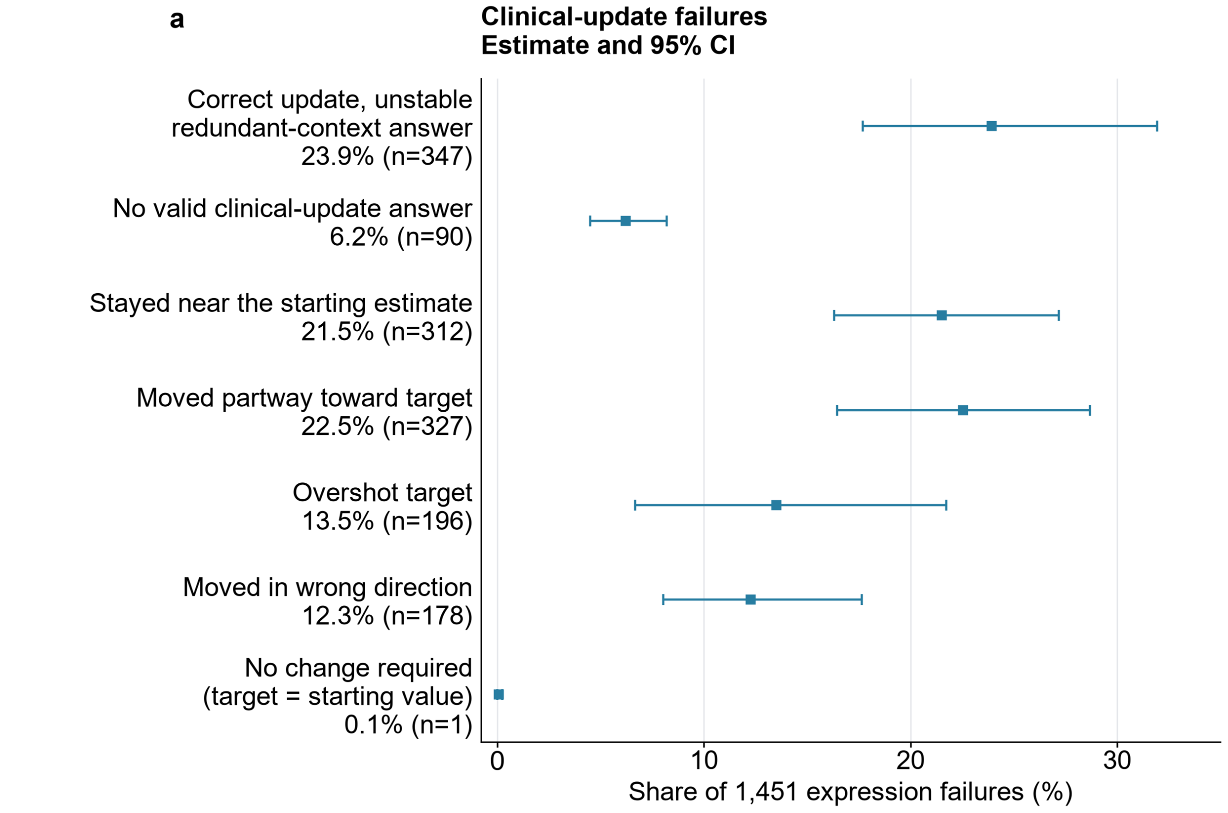

Clinical-update failures
Estimate and 95% CI
Correct update, unstable redundant-context answer 23.9% (n=347)
No valid clinical-update answer 6.2% (n=90)
Stayed near the starting estimate 21.5% (n=312)
Moved partway toward target 22.5% (n=327)
Overshot target 13.5% (n=196)
Moved in wrong direction 12.3% (n=178)
No change required (target = starting value) 0.1% (n=1)
0
10
20
30
Share of 1,451 expression failures (%)


b

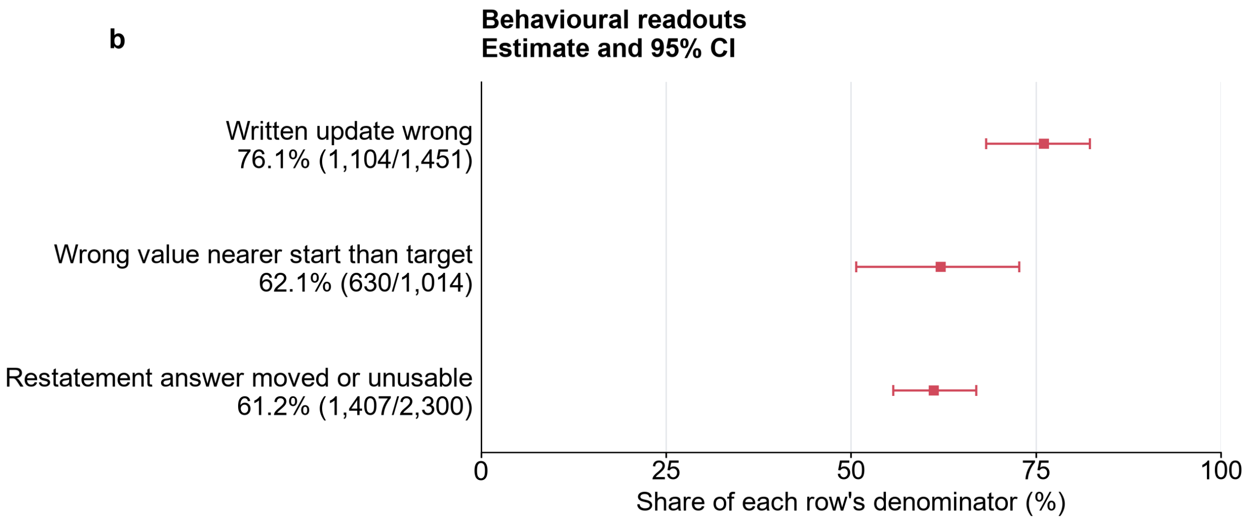

Behavioural readouts
Estimate and 95% CI
Written update wrong 76.1% (1,104/1,451)
Wrong value nearer start than target 62.1% (630/1,014)
Restatement answer moved or unusable 61.2% (1,407/2,300)
0
25
50
75
100
Share of each row's denominator (%)

**Supplementary Figure 23 | Clinical wording failures revealed anchoring on the starting estimate and answer instability.** **a,** Mutually exclusive outcomes among the 1,451 eligible comparisons in which the inputs + calculation arm was correct but a correct clinical update with a stable redundant-context answer was absent (expression failures; Methods). Each point shows a share with its 95% parent-source-cluster bootstrap CI and names the behaviour: answers that gave no valid written number, stayed near the model's starting estimate (the base value, in Methods terms), moved partway toward or overshot the update target, moved in the wrong direction, or required no change because the update target equalled the starting value. "Correct update, but unstable redundant-context answer" identifies the 347 comparisons whose written update was correct but whose redundant-restatement answer (unchanged prevalence restated with an irrelevant detail) moved outside tolerance or became unusable. **b,** Wrong answers stayed near the original estimate, and placebo answers were unstable when prevalence was restated: behavioural readouts with the same interval procedure, each row labelled with its own denominator.

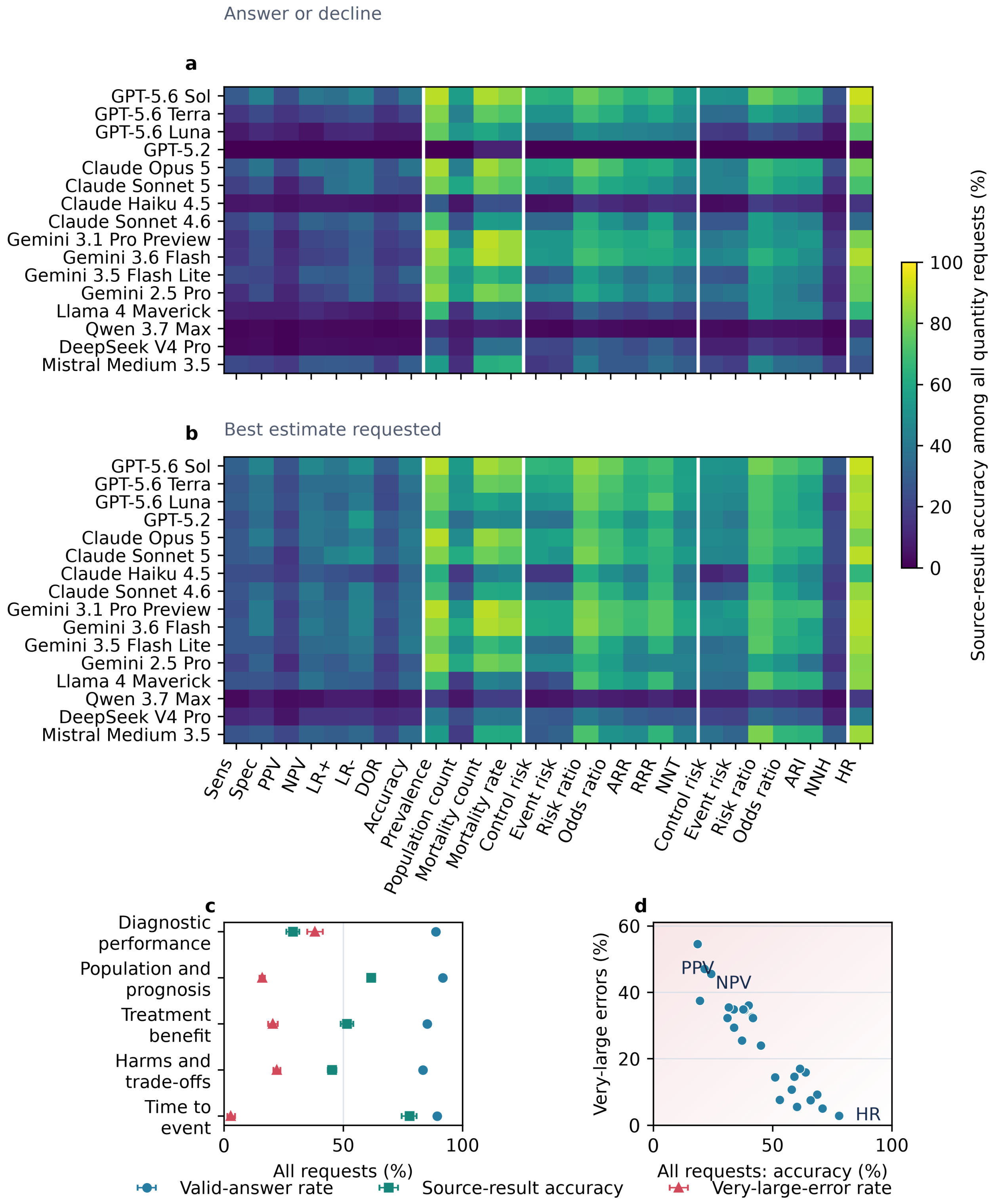

Answer or decline
a
GPT-5.6 Sol
GPT-5.6 Terra
GPT-5.6 Luna
GPT-5.2
Claude Opus 5
Claude Sonnet 5
Claude Haiku 4.5
Claude Sonnet 4.6
Gemini 3.1 Pro Preview
Gemini 3.6 Flash
Gemini 3.5 Flash Lite
Gemini 2.5 Pro
Llama 4 Maverick
Qwen 3.7 Max
DeepSeek V4 Pro
Mistral Medium 3.5
b
Best estimate requested
Sens
Spec
PPV
NPV
LR+
LR-
DOR
Accuracy
Prevalence
Population count
Mortality count
Mortality rate
Control risk
Event risk
Risk ratio
Odds ratio
ARR
RRR
NNT
Control risk
Event risk
Risk ratio
Odds ratio
ARI
NNH
HR
Source-result accuracy among all quantity requests (%)
0
20
40
60
80
100
c
Diagnostic performance
Population and prognosis
Treatment benefit
Harms and trade-offs
Time to event
0
50
100
All requests (%)
d
Very-large errors (%)
0
20
40
60
PPV
NPV
HR
All requests: accuracy (%)
Valid-answer rate
Source-result accuracy
Very-large-error rate

**Supplementary Figure 24 | What numerical results can models reproduce from withheld clinical evidence? a,b,** Source-result accuracy among all quantity requests across 16 model configurations and 26 clinical measures under answer-or-decline (a) and best-estimate (b); vertical boundaries separate diagnostic performance, population/prognosis, treatment benefit, harms/trade-offs and time-to-event measures, and the configuration order was fixed before results and is not a ranking. **c,** Valid-answer rate, source-result accuracy and very-large-error rate as percentages of all quantity requests for each estimation family under best-estimate, with 95% parent-source-cluster bootstrap CIs; circles, squares and triangles denote the three outcomes, respectively. **d,** Source-result accuracy and very-large-error rate by measure under best-estimate; each point is a measure. Higher accuracy and a lower very-large-error rate indicate better performance. The PPV, NPV and hazard-ratio points are labelled; all measure-level values remain in the source data. Abbreviations: Sens, sensitivity; Spec, specificity; PPV and NPV, positive and negative predictive value; LR+ and LR−, positive and negative likelihood ratio; DOR, diagnostic odds ratio; ARR, absolute risk reduction; RRR, relative risk reduction; NNT, number needed to treat; ARI, absolute risk increase; NNH, number needed to harm; HR, hazard ratio. One configuration led four of the five families and trailed the fifth by about one point. The full landscape shows why a single score would obscure large differences among quantities and answer policies.

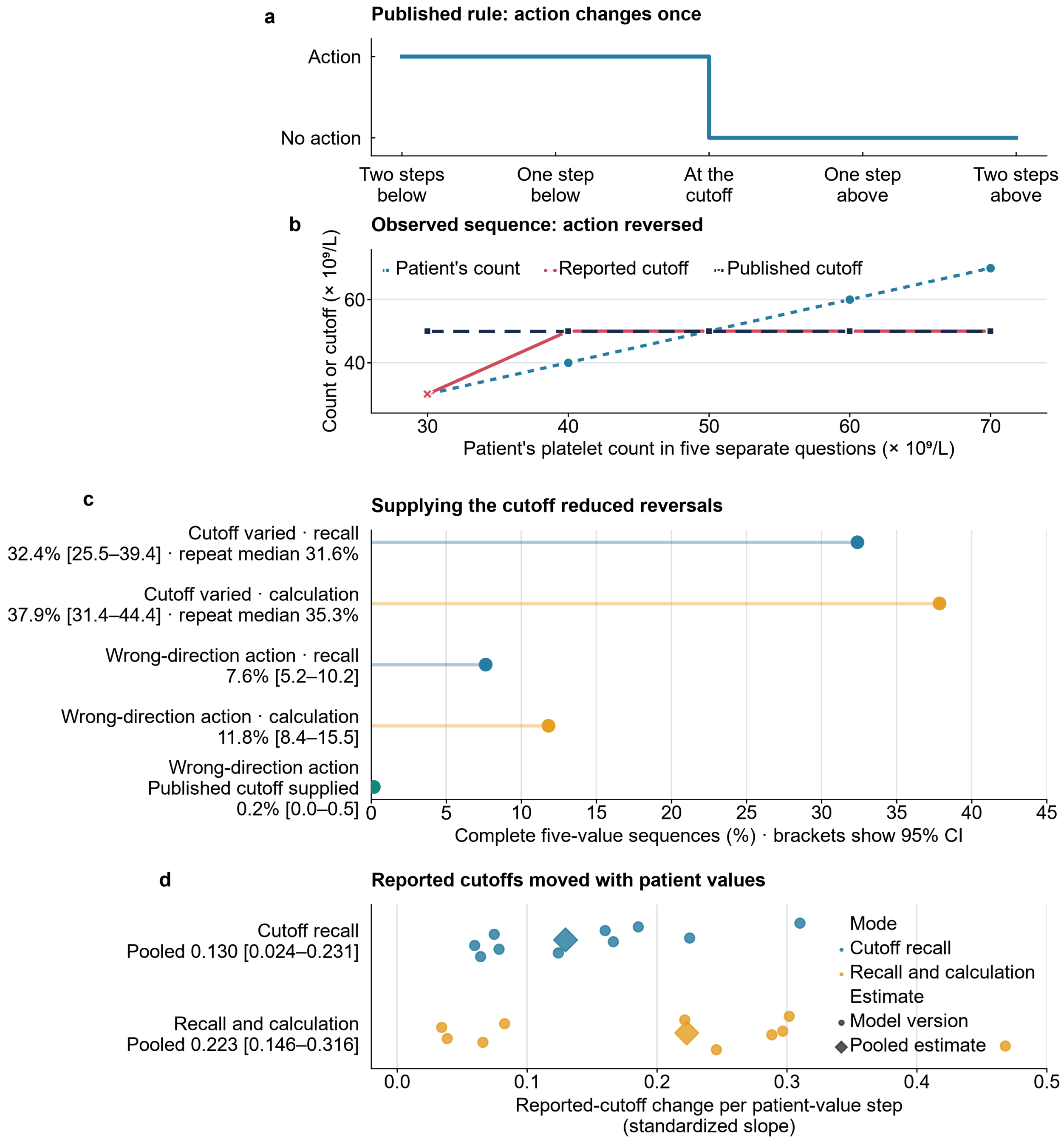
a
Published rule: action changes once
Action
No action
Two steps below
One step below
At the cutoff
One step above
Two steps above
b
Observed sequence: action reversed
Patient's count
Reported cutoff
Published cutoff
Count or cutoff (× 10⁹/L)
60
40
30
40
50
60
70
Patient's platelet count in five separate questions (× 10⁹/L)
c
Supplying the cutoff reduced reversals
Cutoff varied · recall
32.4% [25.5–39.4] · repeat median 31.6%
Cutoff varied · calculation
37.9% [31.4–44.4] · repeat median 35.3%
Wrong-direction action · recall
7.6% [5.2–10.2]
Wrong-direction action · calculation
11.8% [8.4–15.5]
Wrong-direction action
Published cutoff supplied
0.2% [0.0–0.5]
0
5
10
15
20
25
30
35
40
45
Complete five-value sequences (%) · brackets show 95% CI
d
Reported cutoffs moved with patient values
Cutoff recall
Pooled 0.130 [0.024–0.231]
Recall and calculation
Pooled 0.223 [0.146–0.316]
Mode
Cutoff recall
Recall and calculation
Estimate
Model version
Pooled estimate
0.0
0.1
0.2
0.3
0.4
0.5
Reported-cutoff change per patient-value step
(standardized slope)

**Supplementary Figure 25 | An unstable clinical rule reversed the indicated action, and supplying the rule repaired it.** **a,** Five otherwise identical questions place the patient value around a published cutoff (dashed), from two prespecified steps below to two above. The correct action changes at most once, in the source-defined direction. **b,** One observed sequence (platelet transfusion in active upper gastrointestinal bleeding; NICE CG141 cutoff $50 \times 10^9$/L): at patient counts of 30, 40, 50, 60 and $70 \times 10^9$/L the reported cutoff ran 30, 50, 50, 50 and 50, and the inferred action reversed in the wrong direction as the count rose. **c,** Rates over complete five-value sequences, averaged across the 50 recommendations with 95% CIs from resampling recommendations: cutoff variation, 32.4% (cutoff recall) and 37.9% (recall and calculation); repeat-median variation, 31.6% and 35.3%; wrong-direction action reversal, 7.6% and 11.8%, against 0.2% when the published cutoff was supplied. **d,** The reported cutoff moved with the patient's value: standardized slope, 0.130 cutoff-error steps per prespecified patient step under cutoff recall (95% CI, 0.024–0.231) and 0.223 under recall and calculation (95% CI, 0.146–0.316), linking rule instability with patient-level adjustment. Details: Supplementary Methods section 6.
